# A unified 12-lead ECG-language model for interpretation and clinical-endpoint prediction

**DOI:** 10.64898/2026.07.22.26358591

**Authors:** Rohan Banerjee, Juliette Beaulieu, Nicolas Dostie, Blandine Mondésert, Ram Ahuja, Alexis Nolin-Lapalme, Gilbert Jabbour, Shreya Shree Srikanth, Guillaume Marquis-Gravel, Olivier Tastet, Achille Sowa, Julia Cadrin-Tourigny, Jacques Delfrate, Robert Avram

**Affiliations:** Department of Medicine, Montreal Heart Institute, 5000 Belanger Street, QC, H1T 1C8 Canada; HeartWise.ai Laboratory, 5000 Belanger Street, QC, H1T 1C8 Canada; Université de Montréal, Montreal, Quebec, Canada; Karen and Murray Dalfen Arrhythmia Center, Montreal Heart Institute, 5000 Belanger Street, QC, H1T 1C8 Canada; Adult Congenital Heart Disease Center, Montreal Heart Institute, 5000 Belanger Street, QC, H1T 1C8 Canada; Faculty of Medicine, University of Ottawa, Roger Guindon Hall, 451 Smyth Rd, Ottawa, ON K1H 8M5; Mila - Québec AI Institute, Montreal, Quebec, Canada H2S 3H1; Department of Computer Science, University of Toronto, 27 King’s College Cir, Toronto, ON, M5S 1A1 Canada; Cardiovascular Genetics Center, Montreal Heart Institute, 5000 Belanger Street, QC, H1T 1C8 Canada

**Keywords:** Electrocardiography, foundation models, ECG tokenization, large language models, multimodal learning, self-supervised learning, representation learning, clinical endpoint prediction, cardiovascular risk stratification, Canadian Longitudinal Study on Aging, CLSA

## Abstract

Automated electrocardiogram (ECG) interpretation has advanced, yet most systems remain narrow classifiers that emit fixed labels rather than the narratives or endpoint-specific answers clinicians need. Generative approaches could instead produce rich narratives, but are constrained by the gap between continuous biosignals and discrete language tokens. Here we present DeepECG-Tok, which reframes ECG interpretation as a unified instruction-following problem. A residual vector-quantization tokenizer (QINCo) maps 12-lead waveforms to language-model-compatible tokens. Its frozen embeddings achieved a macro-averaged area under the receiver operating characteristic curve (AUROC) of 0.96 for 77-condition classification, outperforming supervised and self-supervised baselines. Aligned with a large language model, a single instruction-tuned model performed ECG interpretation, structured reporting and clinical endpoint prediction, including left ventricular ejection fraction, structural heart disease and atrial fibrillation risk, using 7.27 million question–answer pairs. Frozen-tokenizer diagnostic classification transferred without retraining to two external cohorts, retaining macro-averaged AUROCs of 0.88–0.90, and clinical-endpoint prediction transferred to two further cohorts (external LVEF ≤40% AUROC 0.74–0.76). Evaluated end to end using an ontology-grounded large language model as a judge, which achieved a mean agreement (Cohen’s κ) of 0.82 against two cardiologists, the unified instruction-tuned model scored 0.71 internally and 0.50–0.53 in the same external cohorts. In a blinded reader study, board-certified cardiologists and residents rated its free-text reports comparably to reference clinician reports, with a paired win–tie–loss distribution of 32:35:33 and a forced-choice preference of 0.53 among decided cases, with no significant difference between the model and reference reports in either comparison. These findings establish discrete ECG tokenization as a foundation for general-purpose models that generate clinically useful interpretations and answer diverse questions directly from cardiac waveforms.

## Introduction

Electrocardiography is the most widely used non-invasive diagnostic test in cardiology, supporting the detection of a broad spectrum of cardiovascular conditions^1^. Rising electrocardiogram (ECG) volumes, the specialized training required for expert interpretation, and reported interpretation error rates of roughly 25% even among cardiologists motivate automated interpretation at scale^2^. However, most ECG artificial intelligence (AI) systems remain framed as classification models, constraining their outputs to a small set of predefined diagnostic labels^3–6^. Clinical ECG interpretation is more complex: it integrates rhythm characterization, conduction patterns, waveform morphology and intervals into a structured report. Although modern classifiers can outperform clinicians on narrow supervised tasks^4^, they produce fixed-label predictions rather than structured narratives, targeted explanations or answers to open-ended questions about the ECG. This limitation reflects a broader fragmentation in ECG model development. Beyond reproducing the cardiologist’s interpretation, ECG deep learning can extract digital biomarkers that predict disease unidentifiable by human readers^7–9^. These capabilities have largely been developed in isolation, with separate single-task models for rhythm classification^10^, conduction disorders^11^, ischemia detection^12^, left ventricular ejection fraction (LVEF) estimation^9^, structural heart disease (SHD) prediction^13^, atrial fibrillation (AFib) risk^14^ or report generation^15,16^. Our prior DeepECG self-supervised learning (SSL) foundation model for ECG interpretation reached state-of-the-art performance across 77 diagnostic conditions but still required a separately fine-tuned model for each downstream task, such as LVEF estimation and incident AFib prediction, motivating a single model that serves these clinically distinct tasks and question answering from one shared waveform representation.

Large language models (LLMs) offer a natural interface for such multi-task question and answering, but their application to ECGs is limited by a representation gap between continuous biosignals and discrete language tokens. Bridging this gap requires an ECG representation that compresses raw 12-lead waveforms, preserves diagnostically relevant morphology and can condition a language model across heterogeneous instructions. We adopt Quantization with Implicit Neural Codebooks (QINCo)^17^, an adaptive RVQ framework, to map raw ECG waveforms into compact discrete tokens reusable for both diagnostic representation learning and language-model conditioning.

Here we present DeepECG-Tok, an end-to-end framework that reframes ECG interpretation as a unified instruction-following problem. First, we learn a discrete ECG vocabulary with QINCo that maps raw 12-lead waveforms into language-model-compatible tokens and reusable embeddings. Second, we assemble a multimodal instruction corpus of 7.27 million question-answer (QA) pairs across 19 prompt categories drawn from public and real-world clinical cohorts, including linked clinical endpoints (LVEF, SHD, incident AFib risk, acute coronary occlusion (ACCO) and culprit artery localization) not available in prior ECG-language datasets. Third, we align the frozen ECG tokens with MedGemma 4B-IT through a Q-Former bridge and train a single instruction-following model for free-text interpretation, structured reporting, diagnostic question answering and endpoint prediction. We test generalization without retraining on four external cohorts spanning a population study, an open-access multi-site collection and two echocardiography-linked sets. Because standard text-generation metrics (BLEU^18^, ROUGE^19^, METEOR^20^) capture lexical rather than clinical fidelity, we score outputs with a finding-level framework and benchmark free-text question answering against a panel of cardiologists and residents. Unlike prior ECG-language systems, which deliver either narrative interpretation or isolated clinical endpoints, DeepECG-Tok provides both from one shared waveform representation.

## Methods

This retrospective study was reviewed by the Ethics Board of the Montreal Heart Institute (MHI) (Project #2023-3160) which approved the protocol and waived the need for informed consent as the study represents secondary analysis of data. During manuscript preparation, large language models were used to assist with editorial and stylistic revisions. The authors take full responsibility for the scientific integrity, accuracy, and interpretation of the content presented herein. The external datasets for Medical Information Mart for Intensive Care (MIMIC-IV)^21^, Canadian Longitudinal Study on Aging (CLSA)^22^, CODE-15^23^ and EchoNext-Mini^13^ are available directly from the respective groups.

### Data Source for Model Development

#### Stage 1 and 2 data sources

Raw 12-lead ECG waveforms with corresponding free-text diagnostic reports were obtained from three large-scale datasets: the internal Montreal Heart Institute (MHI) dataset, the public MIMIC-IV-ECG dataset, and the CODE-15 database. MHI and MIMIC-IV provided full diagnostic reports, whereas CODE-15 provided ECG signals without free-text reports. The MHI dataset comprised 1,276,952 12-lead ECGs from 235,423 patients recorded at a specialized quaternary care hospital between 11 April 1997 and 10 November 2023 using the MUSE system (GE HealthCare). The public MIMIC-IV-ECG dataset contributed 708,333 ECGs from 144,441 patients recorded at the Beth Israel Deaconess Medical Center (Boston, USA) between 2008 and 2019, and the CODE-15 dataset contributed 345,797 ECGs from 233,770 patients from the Telehealth Network of Minas Gerais (Brazil) between 2010 and 2017. Data were partitioned at the patient level into training (80%) and test (20%) splits, with patient partitions kept identical to those used for prior DeepECG^3^ foundation model development to enable direct comparison with previously published baselines (Supplementary Fig. 1 and Supplementary Table 1).

Stage 1 tokenizer training used 1.91 million ECGs from the combined MHI, MIMIC-IV and CODE-15 training partitions. Stage 1 used all three cohorts because it optimizes reconstruction only and requires no reports; Stage 2 was restricted to MHI to spare compute, and Stage 3 drew on both MHI and MIMIC-IV, whereas CODE-15 carries no diagnostic reports and was therefore excluded from the text-supervised stages. Stage 2 Q-Former training used 761,386 unique MHI ECGs paired with diagnostic text entries from the MHI training set, yielding 9,565,410 ECG-text training pairs; validation used 94,948 ECG-text pairs from 7,565 held-out MHI ECGs. No patient or waveform assigned to an evaluation split at any stage appeared in any training set. This patient-level disjointness holds across all three stages, so ECGs used only in Stage 1 or Stage 2 training remain disjoint from every evaluation split despite the differing per-stage ECG counts, ensuring strict separation between development and evaluation data across the full pipeline.

#### ECG question-answer dataset construction

For stage 3, we constructed a QA dataset for supervised fine-tuning. The goal was a single unified model capable of performing all downstream tasks within one instruction-following framework, spanning free-text ECG interpretation, structured diagnostic queries, and endpoint prediction. The instruction-tuning corpus comprised 7,274,356 QA pairs generated from 1,087,901 ECGs from 222,684 patients across the MHI (664,697 ECGs from 132,992 patients, 61%) and MIMIC-IV (423,204 ECGs from 89,692 patients, 39%) training splits, spanning 19 prompt categories (Supplementary Tables 2 and 3). For each ECG, prompts were sampled (median 5, interquartile range 4 to 22) across up to five task types: free-text interpretation (35%, including a JSON-structured variant), category-specific diagnostic queries (26%), normal/abnormal classification (20%), demographic and interval queries (15%), and random finding verification (4%). Ground-truth answers were derived from the cardiologist-adjudicated ECG report using a 77-condition diagnostic ontology from our prior work (74 conditions after excluding 3 normal-rhythm labels: sinus rhythm, regular rhythm, and monomorphic QRS morphology) and a BERT classifier that mapped free-text reports to distinct diagnoses and which achieved ≥98% accuracy in this mapping process. For MHI ECGs, five additional endpoint tasks were included: LVEF estimation, SHD detection, incident AFib at 5 years risk prediction, ACCO severity, and culprit-artery identification (Supplementary Table 2). These tasks drew on linked echocardiographic and catheterization data, using the nearest transthoracic echocardiography (TTE) (−90 to +7 days) or coronary angiogram (within 24 hours). Normal ECGs (sinus rhythm without pathology) were capped at 5% of selected waveforms, and minority clinical subgroups were upweighted during training via a weighted sampler without discarding data; validation sets retained natural prevalence distributions.

### Data Pre-processing

All ECG signals were harmonized to a common format prior to analysis (full pipeline in Supplementary Methods S1). Briefly, leads were reordered to a standardized 12-lead sequence, and recordings acquired at 500 Hz (MIMIC-IV, CODE-15) were down-sampled to 250 Hz using a polyphase anti-aliasing filter. Baseline wander was corrected conditionally using a zero-phase 1 Hz high-pass Butterworth filter, applied only when sub-1 Hz spectral energy exceeded the 1–30 Hz band by more than 10-fold. Powerline interference at 50/60 Hz and harmonics was suppressed in the frequency domain using LOESS smoothing, and all amplitudes were rescaled to millivolts.

### Model development

DeepECG-Tok reframes ECG interpretation as instruction following over a shared discrete representation of the waveform, built in three stages (Fig. 1). We first train a self-supervised tokenizer that encodes each 12-lead recording as a compact sequence of discrete codes under a reconstruction objective, producing a reusable ECG vocabulary that is independent of any downstream label. We then align this frozen vocabulary with the embedding space of a pretrained medical language model (MedGemma 4B-IT) through a Q-Former bridge, which compresses each code sequence into a small set of query tokens that condition the language model. Finally, we instruction-tune a single model across heterogeneous ECG tasks, from free-text interpretation and structured reporting to diagnostic question answering and clinical endpoint prediction, holding the tokenizer fixed and adapting the language model through low-rank updates. This yields one model that resolves clinically distinct tasks from a common waveform representation, rather than a separate network fine-tuned for each.

**Figure 1.**
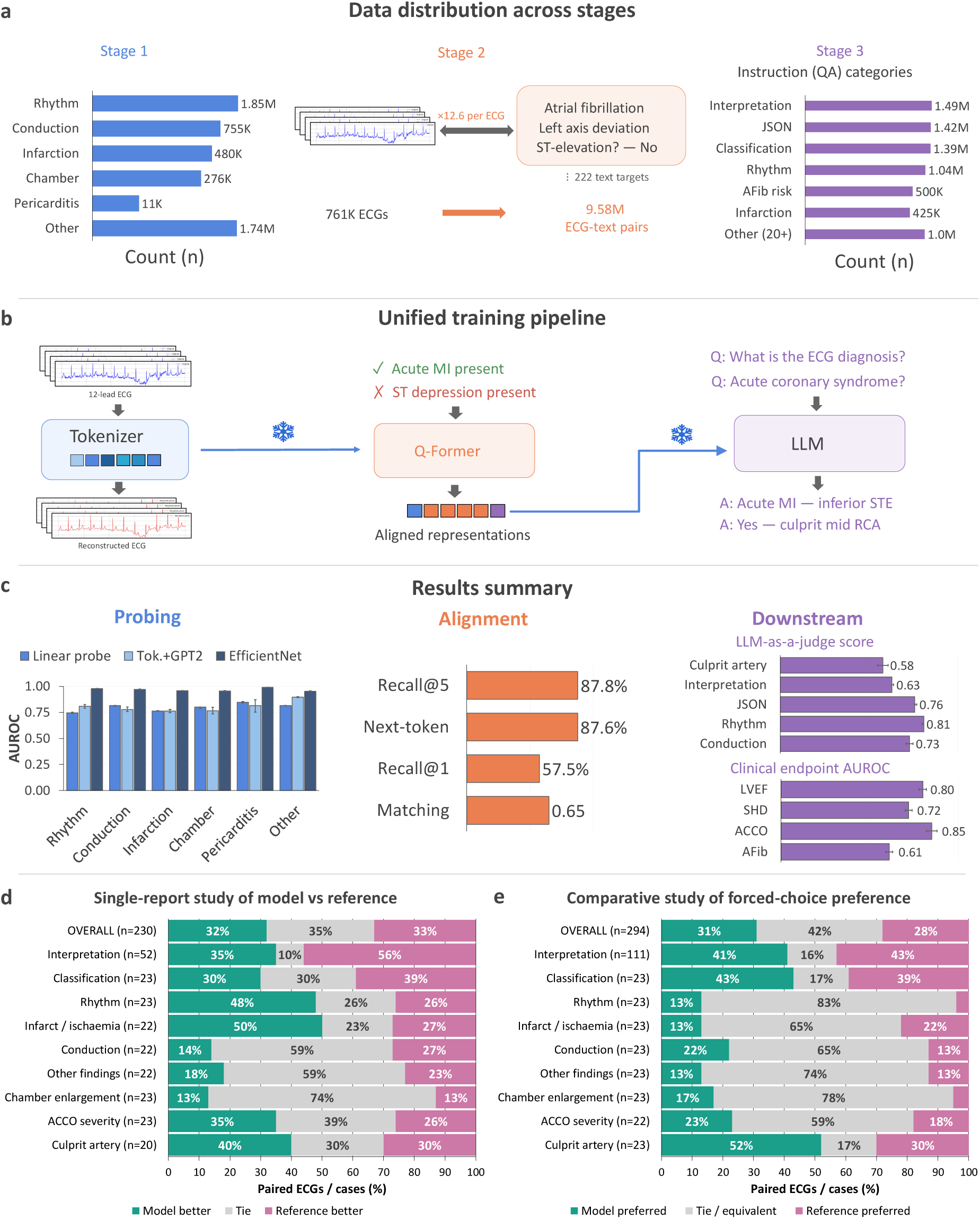
Three-stage training framework and evaluation of DeepECG-Tok. Each panel column corresponds to one training stage: self-supervised ECG tokenization (blue, Stage 1), contrastive ECG-to-text alignment (orange, Stage 2), and instruction tuning (purple, Stage 3). **a,** Data and supervision distribution. Sample counts (n), shown on a logarithmic scale, for the six diagnostic categories represented in the tokenizer training data (Stage 1) and the instruction QA categories used during fine-tuning (Stage 3); the Stage 2 panel summarizes the composition of ECG-text pairs used during alignment. **b,** Unified training pipeline. A 12-lead ECG is encoded into continuous latent vectors and discretized into quantized tokens by the vector-quantized tokenizer; a convolutional decoder reconstructs the original waveform from these tokens during training (Stage 1). Frozen quantized ECG tokens are then aligned to paired free-text reports through a Q-Former bridge (Stage 2). In Stage 3, learned prefix embeddings from the Q-Former condition an instruction-tuned LLM (MedGemma 4B-IT with LoRA adaptation) to generate free-text interpretations and structured clinical outputs in response to natural-language queries. **c,** Results summary. Stage 1 performance is reported as macro-averaged AUROC across six diagnostic categories for three classifier configurations (linear probe, Tok.+GPT-2, EfficientNet). Stage 2 alignment is summarized by ECG-to-report matching AUROC, Recall@1, Recall@5, and next-token prediction accuracy. Stage 3 downstream performance is reported as a category-weighted LLM-as-a-Judge composite score across interpretation categories, as AUROC for four clinical endpoint prediction tasks (LVEF, SHD, ACCO, incident AFib risk), and as the LLM-as-a-Judge category score for culprit artery identification. **d,** Single-report study of model vs reference. Blinded, paired head-to-head comparison in which board-certified cardiologists and trainees rated model-generated or reference (ground-truth, GT) clinician reports independently, without knowing their source. For each category, the 100% stacked bars show the proportion of paired ECGs classified as model better (teal), tie (gray), or GT better (pink). Categories comprise ACCO severity, chamber enlargement, conduction, infarct/ischemia, rhythm, other, classification, culprit artery, and interpretation; OVERALL aggregates all paired ratings (n = 230; model better 32%, tie 35%, GT better 33%). **e,** Comparative study of forced-choice preference. The same readers viewed the model and GT reports for each ECG side by side, in randomized, blinded order, and selected a forced-choice preference. For each category, the 100% stacked bars show the proportion of cases in which the model was preferred (teal), the two reports were rated equivalent (tie, gray), or GT was preferred (pink). Categories are as in panel d; OVERALL aggregates all forced-choice judgments (n = 294; model preferred 31%, tie/equivalent 42%, GT preferred 28%). **Abbreviations:** ACCO: acute coronary occlusion; AFib: atrial fibrillation; AUROC: area under the receiver operating characteristic curve; ECG: electrocardiogram; GT: ground truth; LLM: large language model; LoRA: low-rank adaptation; LVEF: left ventricular ejection fraction; MedGemma: Medical Gemma 4B Instruction-Tuned; QA: question-answer; Q-Former: Query Transformer; SHD: structural heart disease; Tok.: tokenizer; MI: Myocardial Infarction.

Stage 1 trained a self-supervised QINCo residual vector-quantization tokenizer (eight 512-entry codebooks; approximately 0.9M parameters) that compresses each 10-second 12-lead recording approximately 30-fold into 656 discrete codes under a mean-absolute-error reconstruction objective, using 1.91 million ECGs; the tokenizer was frozen for all subsequent stages (Supplementary Methods S3; Supplementary Tables 4 and 5). Stage 2 trained a Q-Former bridge (approximately 199M parameters) to align the frozen ECG codes with the MedGemma 4B-IT embedding space through three objectives, namely ECG-text contrastive learning (ETC), ECG-text matching with hard negatives (ETM), and ECG-conditioned generation (ETG), using 9.57 million ECG-text pairs from 761,386 MHI ECGs (Supplementary Methods S5; Supplementary Table 6). Stage 3 instruction-tuned a single model on 7,274,356 QA pairs, applying low-rank adaptation (LoRA; r = 32) to the top MedGemma layers while jointly training the Q-Former bridge (206.4M trainable of 4.3B total parameters) and holding the tokenizer frozen (Supplementary Methods S6; Supplementary Table 7)

### Model Evaluation

#### Unit of analysis

All internal splits were patient-disjoint. Stage 1 (tokenizer) was trained and evaluated per ECG, with codebook utilization measured per token; Stage 2 (alignment) per ECG and per ECG-report pair; and Stage 3 (instruction tuning) per QA pair, with conventional and LLM-as-a-judge metrics reported per QA pair, the reader study per report and per case, and clinical-endpoint and external-validation metrics per ECG. Denominators are given with each metric in the Results.

### Stage 1 Tokenizer Quality Assessment

Tokenizer reconstruction fidelity was assessed before downstream use. Mean absolute error (MAE) was tracked across training and on the held-out test set to confirm convergence and cross-dataset generalization. A blinded cardiologist reviewed 100 randomly sampled original-versus-reconstructed waveform pairs for preservation of diagnostically relevant morphology (P-wave, QRS configuration, ST-segment contour and T-wave polarity across all 12 leads), with no clinically significant distortion identified.

#### Architecture search and tokenizer selection

We compared different tokenization approaches: ECG-Byte^16^, VQ^24^, RVQ^25^ and QINCo^17^ variants. We also compared sweeping codebook size and utilization to identify a codebook configuration that was both expressive (a large discrete vocabulary with high reconstruction fidelity) and stable (consistently high codebook utilization). In our main analysis, a codebook size of 512 was selected since it matched the reconstruction quality and utilization stability of the best alternative (256) while providing a larger discrete vocabulary (Supplementary Table 5; strategy comparisons in Supplementary Table 4). The tokenizer weights were kept frozen (not trainable) for all subsequent analyses.

#### Probing analyses to verify representation quality

We probed the quantized latent sequences using a linear head and an EfficientNetV2-based nonlinear 1D classifier (the primary analysis), each optimized with multi-label binary cross-entropy across the 77 ECG interpretation conditions spanning six AHA-aligned^26^ diagnostic categories (Supplementary Table 8), with ground truth from a validated BERT-based^27^ report classifier (Supplementary Methods S2). These categories included chamber enlargement, ischemic changes, conduction abnormalities, wave abnormalities (i.e. T wave inversion, etc) and pericarditis. Frozen-tokenizer performance under both probes was evaluated on the MHI and MIMIC-IV test sets against DeepECG-SL and DeepECG-SSL baselines (Supplementary Table 9), and against ECG-GPT^15^ on its published label-assessment protocol using PTB-XL^28^ (Supplementary Methods S9).

#### Report-generation feasibility evaluation

To assess whether the learned discrete token sequence could support autoregressive ECG-to-text generation, lightweight language-model decoders were attached to the frozen tokenizer embeddings. We first evaluated a GPT-2^29^ (124 million parameters) decoder for report generation, then trained a Llama 3.2^30^ (1 billion parameters) decoder on the same tokenizer outputs to enable an architecture-matched comparison with ECG-Byte. Decoder performance was evaluated on the interpretation subset of the combined internal test set, corresponding to the free-text ECG interpretation task defined in the ECG question-answer (QA) dataset construction section. Performance was reported using ROUGE-1, ROUGE-L, BLEU-4 and METEOR, with full comparisons provided in Supplementary Methods S4 and Supplementary Table 10.

### Stage 2. ECG-language alignment

Q-Former alignment was assessed on the held-out MHI validation set (7,565 ECGs) across all three pre-training objectives. ETC was quantified by Recall@K (K = 1, 5, 10, 25) over a per-batch candidate bank of diagnostic text descriptions. ETM was evaluated as binary accuracy over fused query-text representations distinguishing matched from mismatched pairs, with hard negatives drawn from dynamic in-batch mining (the most similar non-matching text per ECG under current representations) and 10 clinically curated confusability families (for example, ST elevation versus depression in the same territory, ventricular tachycardia (VT) versus supraventricular tachycardia (SVT) with aberrancy, LBBB versus ventricular pacing; Supplementary Table 11); binary QA items contributed automatic yes/no counterparts, forcing the bridge to attend to the signal rather than text-only cues. ETG was assessed by teacher-forced next-token prediction on clinician reports tokenized with MedGemma 4B, with greedy-decoded outputs inspected for capture of primary and secondary findings. Tail-class retrieval was characterized on the 50 rarest conditions in the dataset. All metrics were tracked across the 10 training epochs (Supplementary Table 12). Twelve hyperparameters were explored via a Bayesian sweep with Hyperband early stopping (Supplementary Table 6), selecting the configuration with the lowest validation contrastive loss.

### Stage 3. Conventional metrics, LLM-as-a-judge, and human expert adjudication

Model outputs were evaluated across **<u>two</u>** complementary frameworks (Figure 2b; Supplementary Methods S7). **Lexical similarity** used ROUGE-1, ROUGE-2, ROUGE-L, BLEU-4 and METEOR on held-out interpretation samples. Clinical fidelity used a hierarchical **LLM-as-a-judge, in which** structured and numeric outputs were scored deterministically field-by-field against clinical tolerance thresholds (heart rate ±5 bpm, PR/QT ±20 ms, LVEF ±5%; Supplementary Table 13), while free-text narratives were scored by MiniMax-M2.5^31^ (temperature = 0) prompted to reason as a board-certified cardiologist would and supported by an 847-term ECG ontology designed by 3 cardiologists for synonym resolution. Findings were classified as identified, missed, or hallucinated and scored as |identified| / (|identified| + |missed| + |hallucinations|), aggregated as weighted averages across 19 categories (Supplementary Table 13). To validate the judge, two board-certified cardiologists independently re-scored a random sample of 500 QA answers using this same rubric (rating each question-and-answer on the 0-to-1 identified/(identified + missed + hallucinated) scale while viewing the corresponding ECG), and their ratings were compared with the LLM-as-a-judge scores, yielding a mean Cohen’s κ of 0.82 (Supplementary Methods S7).

**Figure 2.**
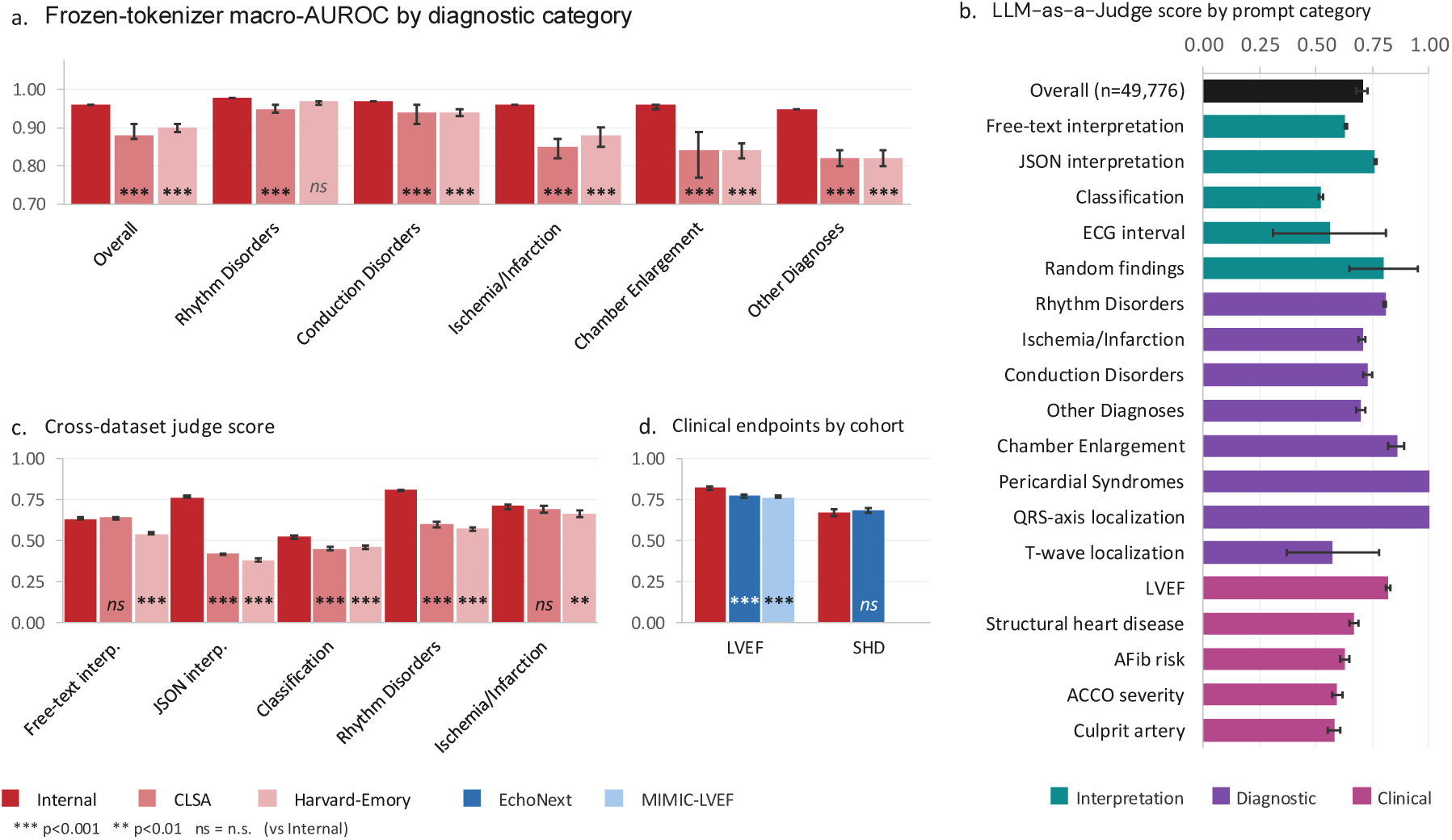
External validation and cross-cohort generalization of DeepECG-Tok across diagnostic classification, ECG interpretation, and clinical-endpoint prediction. a, Multi-label classification performance of frozen tokenizer embeddings, measured as macro-averaged AUROC using an EfficientNetV2 classifier, shown by diagnostic category (Overall, Rhythm Disorders, Conduction Disorders, Ischemia/Infarction, Chamber Enlargement and Other Diagnoses) for the internal (MHI, n = 416,467), CLSA (n = 5,079) and Harvard-Emory (n = 5,100) test sets; asterisks denote a two-sided bootstrap difference test (1,000 iterations) comparing each external cohort with the internal cohort. b, LLM-as-a-Judge evaluation scores for the internal validation cohort (MHI + MIMIC, n = 49,776 question-answer pairs) across 19 prompt categories, ordered by sample size and grouped by task type (interpretation tasks, diagnostic categories and clinical endpoints); the overall score represents the category-weighted aggregate computed using predefined clinical task weights and is shown in black. c, Cross-dataset comparison of LLM-as-a-Judge scores across the five evaluation categories common to all cohorts, for the internal (n = 49,776), CLSA (n = 20,289) and Harvard-Emory (n = 20,400) cohorts; asterisks denote two-sided Mann-Whitney U tests comparing each external cohort with the internal cohort. d, LLM-as-a-Judge scores for the clinical-endpoint tasks, left ventricular ejection fraction (LVEF) and structural heart disease (SHD), comparing the internal MHI cohort with the external EchoNext (LVEF and SHD) and MIMIC-LVEF (LVEF only) cohorts; asterisks denote two-sided Mann-Whitney U tests comparing each external cohort with the internal cohort. Inline bars show point estimates with bootstrap 95% confidence interval (CI) whiskers, and all error bars represent bootstrap 95% CIs from 1,000 iterations with seed 42. Across all panels, comparisons are made relative to the internal MHI cohort, with significance levels indicated as *\*\*p* < 0.01, *\*\*\*p* < 0.001 and ns, not significant. **Abbreviations**: MHI: Montreal Heart Institute; MIMIC: Medical Information Mart for Intensive Care; CLSA: Canadian Longitudinal Study on Aging; LVEF: left ventricular ejection fraction; SHD: structural heart disease; AUROC: area under the receiver operating characteristic curve; LLM: large language model.

Furthermore, we performed a blinded, randomized, fully-crossed multi-clinician multi-case study of free-text interpretation of AI generated answers against the paired reference report (cardiologist text), reported per TRIPOD-LLM^32^. We annotated free-text ECG interpretations, not the full dataset. All ECGs used in the reader study were drawn from the held-out test set and were excluded from all three training stages, so no reader-study patient contributed to tokenizer, alignment, or instruction-tuning data. Six readers (three attending cardiologists, two of them electrophysiologists, and three residents) rated each report blinded to its source (AI or cardiologist) and against the ECG tracing itself. Each report was scored on three ordinal 1-4 axes: correctness, completeness, and clarity (Mayo-inspired schema). Readers also gave an overall grade, flagged errors/hallucinations with their type and clinical-harm severity, and judged primary-diagnoses correctness. The aggregate score was the mean of the three axes.

Two annotation designs were used on independent, identically-stratified samples:

- Dataset A (single-report absolute rating): each report shown alone. 847 ratings collected across 472 reports, mean 1.8 of 3 target ratings per report.
- Dataset B (forced-choice comparison): both reports shown side by side in randomized order. 294 forced-choice judgments across 237 cases.

Target was up to three labels per report. Win/tie/loss was derived per ECG from the Dataset A aggregates (model win if its aggregate exceeded the reference, tie if equal, loss otherwise) and tested by paired Wilcoxon signed-rank and McNemar; the preference rate was the share of Dataset B cases selecting the model. Both, with the absolute aggregate Likert quality for model and reference, are reported overall, per category and per reader, with Wilson intervals against 50% and bootstrap CIs, ties reported explicitly. Non-inferiority was pre-specified at a - 0.05 absolute correctness margin (one-sided α = 0.025), with reader-experience tier and source dataset as pre-planned subgroups and inter- and intra-reader reliability by Gwet’s AC1/AC2 on the dichotomized ratings.

Finally, to confirm outputs were conditioned on ECG content rather than text priors, we re-ran inference on 996 stratified MHI test QA pairs under three input conditions with identical prompts and parameters: the correctly preprocessed ECG (normal), an all-zeros tensor (zeroed), and a different patient’s ECG (shuffled). Conditions were compared on LLM-as-a-judge scores, lexical metrics (ROUGE-L, BLEU-4, METEOR), cross-condition output similarity (exact-match rate and pairwise ROUGE-L), and signal-absence detection rate (proportion of outputs with explicit refusals such as “cannot analyze”), with paired per-example win/tie/loss of normal versus each alternative (Supplementary Methods S8).

### External Validation

External validation was performed on four independent cohorts without retraining or adaptation, selected to assess generalization across populations, clinical settings, and prediction tasks (Supplementary Table 14); the distribution of evaluation QA pairs across prompt categories for each cohort is reported in Supplementary Table 15. For ECG interpretation and multi-label classification, we evaluated two cohorts with free-text ECG reports. The CLSA^22^, a national population-based cohort of community-dwelling adults aged 45 to 85 years, contributed 5,079 ECGs from 5,079 patients drawn from 29,427 baseline recordings. The Harvard-Emory ECG Database ^33^, an open-access collection of 12-lead ECGs acquired during routine care at Massachusetts General Hospital and Emory University Hospital, contributed 5,100 ECGs from 5,100 patients drawn from 998,844 recordings. Both cohorts were selected by multi-label stratified sampling (seed 42) across pathological categories, capping normal tracings at 5% to enrich for clinically relevant findings. For clinical endpoint prediction, no additional sampling was applied: we used all MIMIC-IV ECGs with a linked echocardiographic LVEF measurement (5,134 ECGs, 1,146 patients) and the published EchoNext^34^ held-out test set (5,442 ECGs, 5,442 patients, one ECG per patient; LVEF available for 4,827), comprising 12-lead ECGs from patients referred for echocardiography at Columbia University Irving Medical Center. All external waveforms were harmonized with the same preprocessing pipeline used during training (Supplementary Methods S1).

### Statistical Analysis

Three pre-specified primary endpoints were applied identically to internal and external cohorts: macro-averaged classification AUROC cohort for any binary classification task; heart rate and LVEF correlation by MAE and Pearson *r*; and free-text interpretation fidelity under the LLM-as-a-judge framework (Supplementary Methods S7). Secondary endpoints characterized generalization and verified each stage: per-category AUROC, report-generation lexical similarity (ROUGE-1, ROUGE-2, ROUGE-L, BLEU-4, METEOR), ICC(2,1), and the signal-dependency analysis for the final model. For Stage 1, we assessed reconstruction fidelity, codebook utilization, and probing AUROC across the 77-condition ontology. For Stage 2, we assessed Recall@K, ECG-text matching accuracy, teacher-forced generation, and tail-class retrieval on the 50 rarest conditions. Representation quality was benchmarked against DeepECG-SL and DeepECG-SSL on the identical patient-disjoint internal test set. Tokenizer report generation was benchmarked against ECG-Byte^16^ under a matched Llama 3.2-1B backbone and LoRA budget, and interpretation against ECG-GPT’s^15^ published protocol (Supplementary Methods S9). Binary clinical endpoints were scored from generative output: LVEF thresholds (≤40%, <50%) from the parsed numeric prediction, and yes/no biomarkers (5-year incident AFib risk, ACCO, SHD) from the renormalized first-token P(Yes). LVEF endpoints were analyzed complete-case: ECGs without a linked echocardiographic LVEF were excluded from the LVEF analysis (in the EchoNext held-out test set, 4,827 of 5,442 ECGs had an available LVEF). AUROC, AUPRC, sensitivity, specificity, PPV, and NPV were then computed at a validation-set Youden threshold against linked ground truth. All 95% confidence intervals were bootstrapped (1,000 iterations, seed = 42), stratified by diagnostic class for classification; between-cohort AUROC and judge-score differences were tested by two-sided bootstrap and Mann-Whitney U, respectively.

Fairness was assessed at two stages, with subgroups by sex and age band (below 55, 55 to 75, above 75 years). For the Stage 1 classifier we applied equalized odds per-condition predictions^35^ were binarized at each label’s threshold, TPR and FPR micro-averaged across the 77 labels per subgroup, and disparity summarized as ΔTPR and ΔFPR. No consensus acceptability threshold for equalized-odds disparity exists in clinical AI, so we pre-specified an absolute tolerance of 0.10 for ΔTPR and ΔFPR as a descriptive screening bar rather than a validated criterion, and report all subgroup gaps regardless of whether they cross it. As equalized odds requires a threshold classifier, Stage 3 fairness was assessed as parity of the LLM-as-a-judge composite across the same subgroups, tested by two-sided Mann-Whitney U with female sex and age below 55 as references. Given the large pooled sample (n = 49,776), we interpreted Stage 3 fairness primarily from effect sizes and composite-difference CIs rather than p values. Because equalized odds is prevalence-sensitive and the base rates of several conditions differ genuinely by age and sex, these metrics are reported as descriptive parity and not as evidence of an absence of bias. All estimates carried 95% bootstrap CIs (1,000 resamples of 80% of dataset, seed = 42). Analyses were restricted to the internal test set.

## Results

The combined training cohort comprised 1,914,615 ECGs from 530,519 patients across MHI, MIMIC-IV, and CODE-15, with mean ages of 51.0 ± 19.9 years to 60.1 ± 16.2 years and female proportions of 44.1%, 51.8%, and 59.4%, respectively (Table 1). External validation was performed on four independent cohorts totalling 20,755 ECGs from 16,767 patients: CLSA, Harvard-Emory, MIMIC-IV echocardiography subset, and EchoNext.

**Table 1.** Baseline characteristics of the training cohort. Patient-level characteristics for each dataset included in tokenizer and language model training. Data from the MHI, MIMIC-IV, and CODE-15 are shown separately. Exams per patient are reported as median [interquartile range], age per patient as mean ± standard deviation, and sex as n (%). **Abbreviations:** MHI: Montreal Heart Institute; MIMIC: Medical Information Mart for Intensive Care; CODE-15: Clinical Outcomes in Digital Electrocardiology; SD: standard deviation; IQR: interquartile range; Hz: Hertz. Counts reflect the patient-level training partition (80% split); full-cohort totals are reported in the Methods.

|  | MHI | MIMIC-IV | CODE-15 |
| --- | --- | --- | --- |
| **Number of ECGs** | 1,017,719 | 551,099 | 345,797 |
| **Number of patients** | 184,210 | 112,539 | 233,770 |
| **Exams per patient, Median [IQR]** | 2 [1 to 6] | 2 [1 to 5] | 1 [1 to 2] |
| **Mean age per patient, years** | 60.1 ± 16.2 | 57.9 ± 19.5 | 51.0 ± 19.9 |
| **Male, n (%)** | 102,903 (55.9%) | 53,031 (47.1%) | 94,807 (40.6%) |
| **Female, n (%)** | 81,307 (44.1%) | 58,308 (51.8%) | 138,963 (59.4%) |
| **Sampling frequency** | 250 Hz | 500 Hz (resampled to 250 Hz) | 500 Hz (resampled to 250 Hz) |

### Stage 1

QINCo achieved the lowest held-out reconstruction error of the three quantizers evaluated (test mean absolute error 0.03, versus 0.04 for RVQ and 0.07 for vanilla VQ) while maintaining complete codebook usage, and a 512-code codebook was selected for all downstream analyses (Supplementary Tables 4 and 5; Supplementary Figs. 2–4).

The frozen tokenizer preserved diagnostically relevant information under both evaluation modes. On the 77-label ECG interpretation classification task (held-out internal test set, n = 416,467), probing of its embeddings reached a macro-AUROC of 0.79 (0.78–0.79) with a linear head and 0.96 (0.95–0.96) with an EfficientNetV2 classifier (Figure 1c; Supplementary Table 9), exceeding DeepECG-SL (0.94, 0.94–0.94) and DeepECG-SSL (0.93, 0.93–0.94) (Supplementary Table 16) and matching or surpassing ECG-GPT on its published protocol (Supplementary Table 17). Attaching a Llama 3.2-1B decoder to the same frozen embeddings, fine-tuned via LoRA (rank 16, applied to query and value projections) on 1.49 million ECG-report pairs, enabled generative interpretation (n = 10,000 evaluation ECGs) at a macro-AUROC of 0.72 (0.71–0.73), versus 0.50 (0.50–0.51) for ECG-Byte with the same backbone (Supplementary Table 10; Supplementary Methods S4, S9).

### Stage 2

Using these frozen tokenizer embeddings, ECG-language alignment on the held-out MHI validation set (7,565 ECGs) achieved ETC Recall@1 of 0.58 (95% CI, 0.56 to 0.59) and Recall@5 of 0.88 (95% CI, 0.87 to 0.89) by epoch 10, ETM AUROC of 0.65 (large-sample 95% CI, 0.64 to 0.66), and ETG next-token prediction accuracy of 87.6%. Tail-class retrieval on the 50 rarest diagnostic strings reached Recall@5 of 0.11 (95% CI, 0.09 to 0.13), an expected stress test under extreme class imbalance in the many-to-many ECG-text setting, where each ECG can have multiple valid matched strings and rare strings are more likely to rank below common matched findings (Supplementary Table 12).

### Stage 3

Following LoRA-based instruction tuning, the model was evaluated on 49,776 question-answer pairs (24,995 MHI; 24,781 MIMIC-IV) across 19 prompt categories using the LLM-as-a-judge framework (Figure 2b; Supplementary Table 3). The framework, scores and rationale were validated against two board-certified cardiologist adjudications on 500 randomly sampled examples which achieved a strong average correlation of LLM-score vs cardiologist’s score (average Cohen’s kappa of 0.82; Supplementary Table 18). Overall judge performance yielded a weighted composite score of 0.71 (95% CI, 0.68 to 0.73). Structured JavaScript Object Notation (JSON) interpretation outscored free-text (0.76 versus 0.63; Supplementary Table 19). Representative scored model outputs, with judge rationales across diagnostic categories, are provided in Supplementary Table 20.

On ECG-GPT’s published six-label evaluation protocol, instruction-tuned DeepECG-Tok was benchmarked against ECG-GPT (AUROC, DeepECG-Tok versus ECG-GPT): AFib 0.92 versus 0.96, sinus tachycardia 0.96 versus 0.95, sinus bradycardia 0.91 versus 0.91, complete LBBB 0.95 versus 0.97, complete RBBB 0.96 versus 0.98, and first-degree atrioventricular block 0.66 versus 0.85 (Supplementary Table 21; protocol detailed in Supplementary Methods S9).

For continuous cardiac parameters, DeepECG-Tok showed strong correlation with measured values for heart rate (MAE 2.21 bpm; Pearson *r* 0.93, 95% CI 0.92 to 0.94; ICC 0.96). For LVEF, correlation was moderate (MAE 8.41%; *r* 0.56; ICC 0.68), with an AUROC of 0.80 for detecting LVEF ≤40% (sensitivity 0.66 internally, 0.46 to 0.48 externally; PPV 0.48 internally, 0.46 to 0.54 externally; Table 2; Supplementary Table 22). Structural heart disease, five-year incident AFib, and acute coronary occlusion detection are summarized in Figure 3 and Table 2 (judge score 0.67; AUROC 0.61; AUROC 0.85, respectively). In the MHI test cohort, the five-year incident-AFib endpoint comprised 1,160 events among 3,121 labeled ECGs (37.2%), and the two-year endpoint 1,069 events (34.3%).

**Figure 3.**
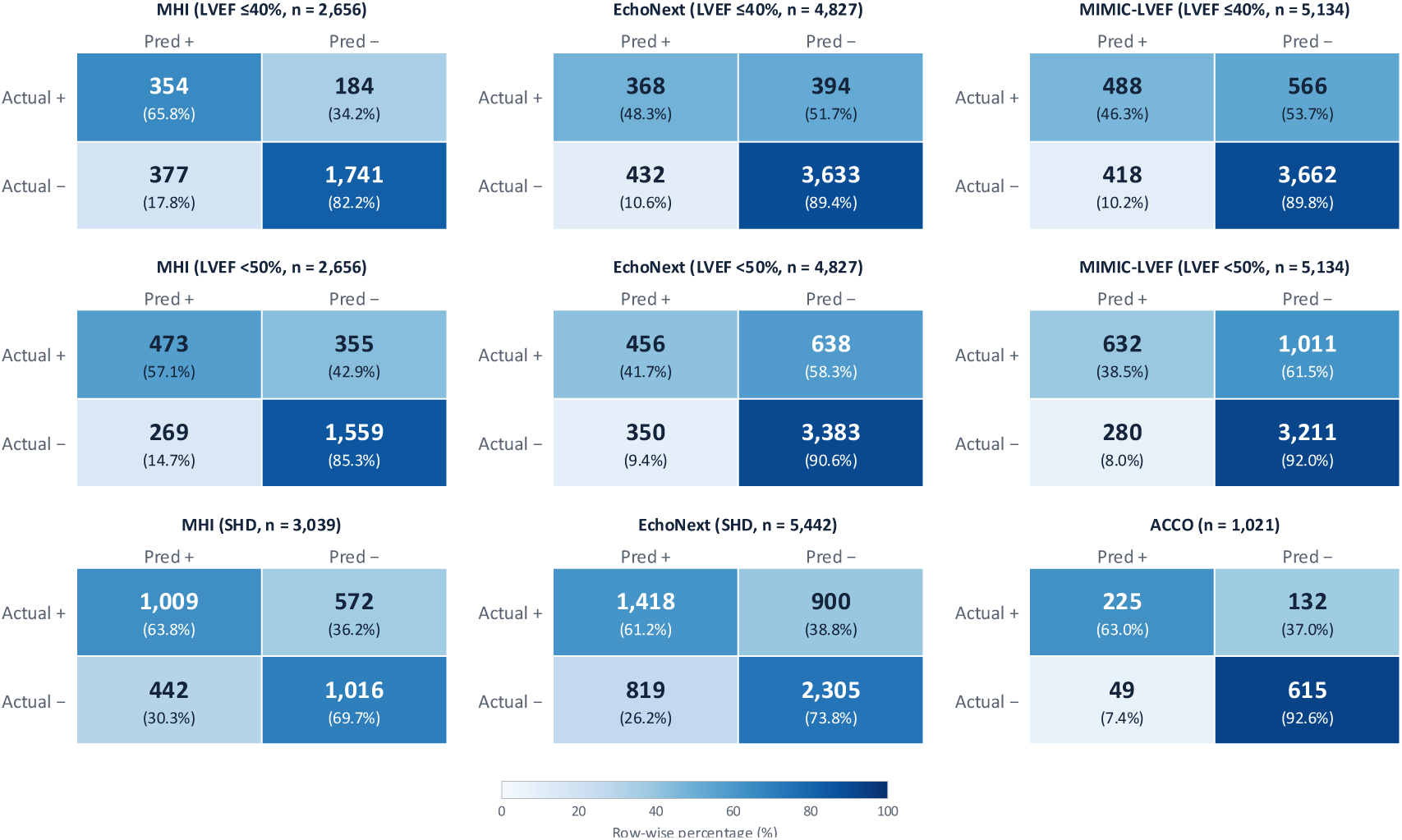
Binary classification performance across cardiac endpoints and external cohorts. Confusion matrices for four tasks derived from model-generated reports: left ventricular ejection fraction (LVEF) ≤40%, LVEF <50%, structural heart disease (SHD; present vs absent), and acute coronary occlusion (ACCO; present vs absent), with each panel labeled by task and cohort. Each matrix displays absolute counts with row-wise percentages (sensitivity for the positive row, specificity for the negative row) in parentheses. Color intensity reflects the proportion of each true-label row assigned to each predicted category (0 to 100%). Internal validation was performed on the MHI held-out test set; external validation was performed on MIMIC-LVEF (n = 5,134 ECGs from 1,146 patients) and EchoNext (n = 4,827 ECGs for LVEF; n = 5,442 for SHD). SHD was evaluated on MHI and EchoNext only, as linked echocardiographic labels were unavailable in MIMIC-LVEF. ACCO was evaluated on MHI (n = 1,021 ECGs), with sensitivity 63.0% (225/357) and specificity 92.6% (615/664). Ground-truth LVEF values were derived from echocardiograms linked to ECGs within each cohort; SHD and ACCO labels were defined as described in Methods. **Abbreviations: ACCO:** acute coronary occlusion; LVEF: left ventricular ejection fraction; SHD: structural heart disease; MHI: Montreal Heart Institute; MIMIC: Medical Information Mart for Intensive Care.

**Table 2.**
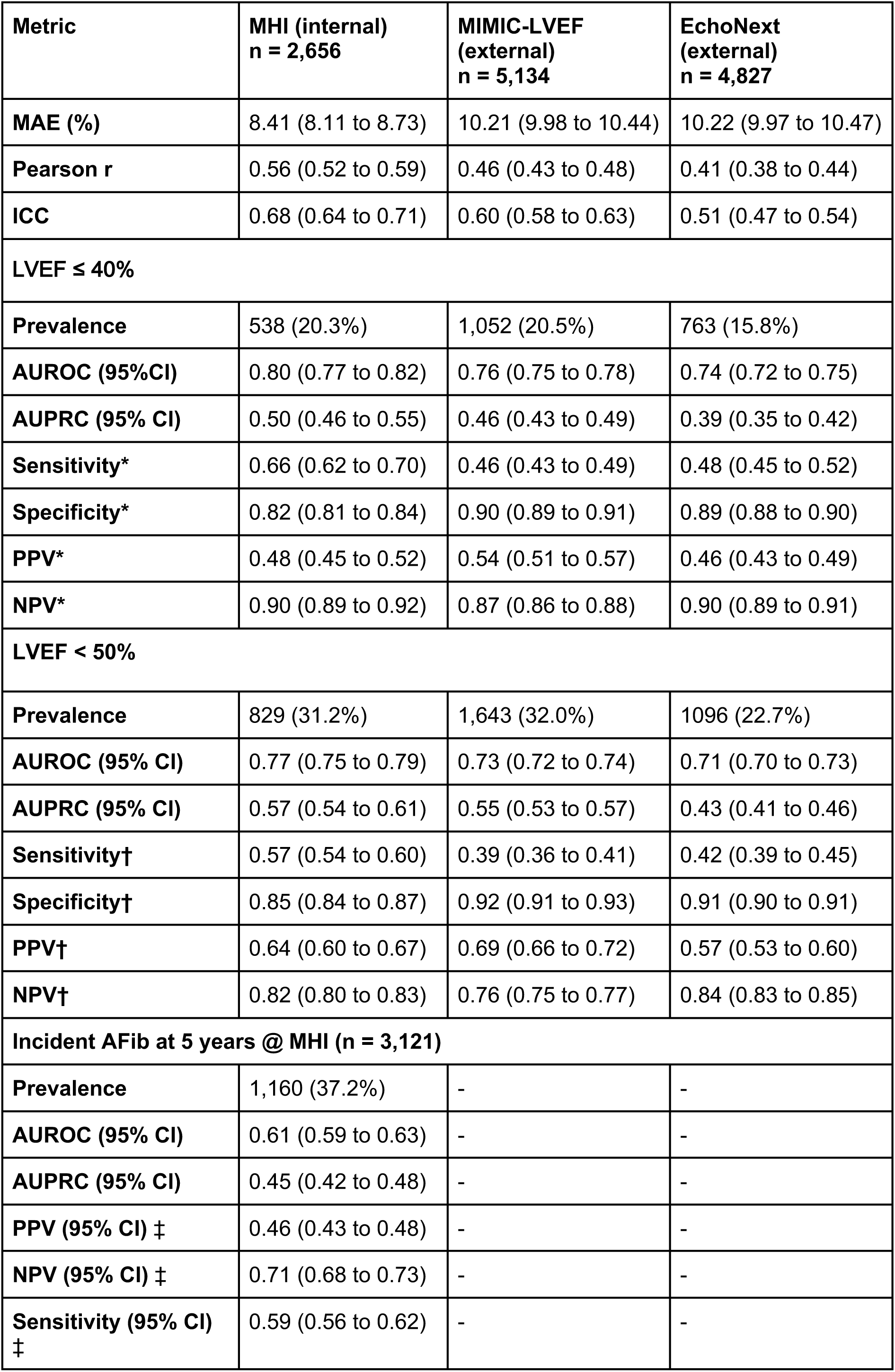
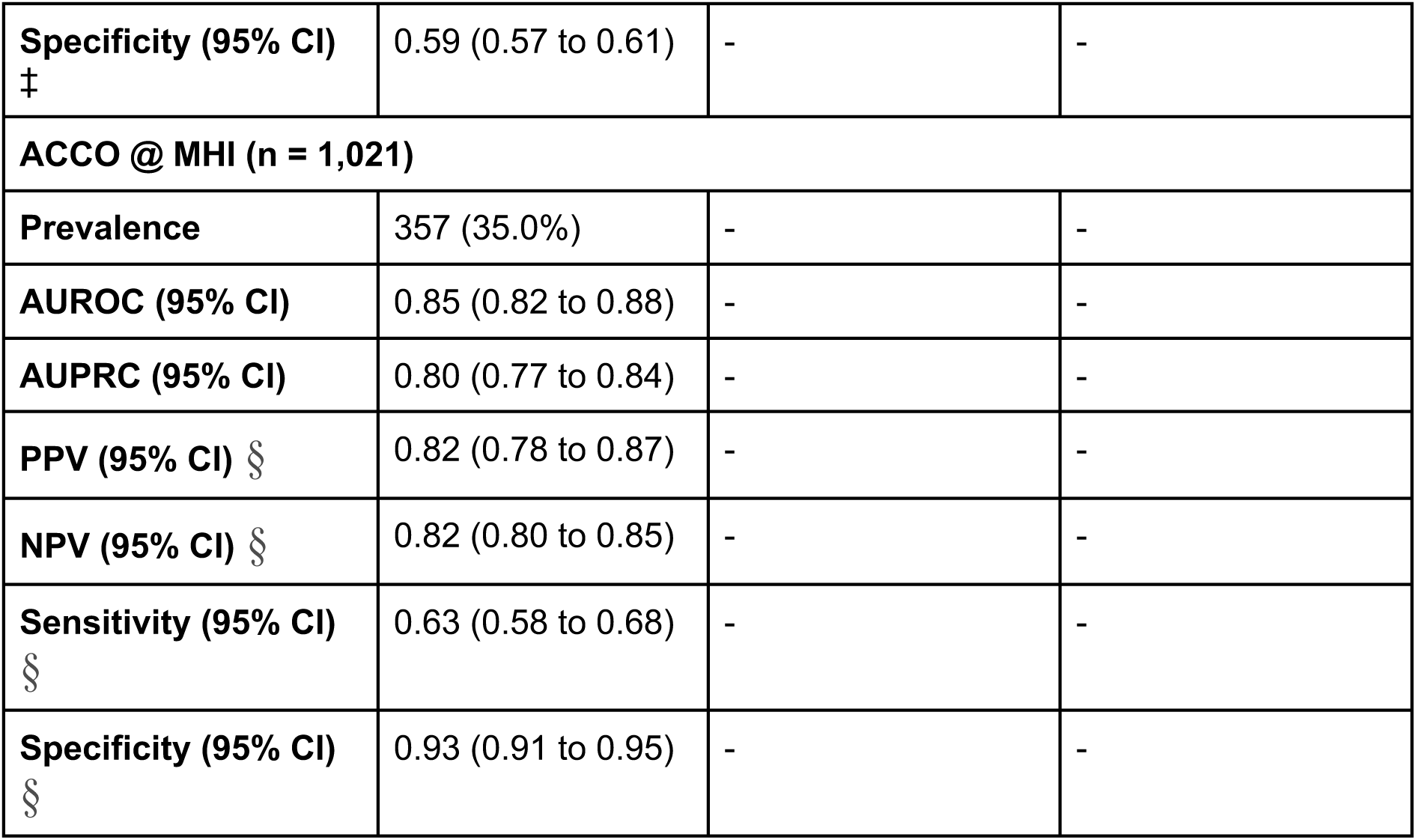
Agreement and classification performance for model-predicted and echocardiography-derived LVEF across internal and external validation cohorts. MAE, Pearson correlation coefficient r, and ICC (2,1) were computed between numeric LVEF values extracted from model-generated reports and ground-truth LVEF from linked echocardiographic measurements. AUROC, AUPRC, sensitivity, specificity, PPV, and NPV were computed at two prespecified clinical thresholds, LVEF ≤40% (moderately-to-severely reduced) and LVEF <50% (any reduction). AUROC and AUPRC used the negative predicted LVEF as the discrimination score; sensitivity, specificity, PPV, and NPV were calculated by binarizing the model-predicted LVEF at the same clinical threshold as the ground-truth label, applied identically across all cohorts. Outcome prevalence (MHI, MIMIC-LVEF, EchoNext) was 20.3%, 20.5%, and 15.8% for LVEF ≤40% and 31.2%, 32.0%, and 22.7% for LVEF <50%. MHI served as internal validation (the model was trained on MHI LVEF labels). MIMIC-LVEF and EchoNext served as external validation (no LVEF from either cohort was used during training). All values are reported with 95% confidence intervals from 1,000 bootstrap iterations, 80% sample (seed = 42). Abbreviations: LVEF: left ventricular ejection fraction; MAE: mean absolute error; ICC: intraclass correlation coefficient; AUROC: area under the receiver operating characteristic curve; AUPRC: area under the precision-recall curve; PPV: positive predictive value; NPV: negative predictive value; AFib: Atrial fibrillation; ACCO: Acute coronary occlusion. * Threshold = 0.45; †: Threshold = 0.45; ‡ Threshold = 0.992; § Threshold = 0.119

| **Metric** | **MHI (internal)**<br>**n = 2,656** | **MIMIC-LVEF**<br>**(external)**<br>**n = 5,134** | **EchoNext**<br>**(external)**<br>**n = 4,827** |
| --- | --- | --- | --- |
| **MAE (%)** | 8.41 (8.11 to 8.73) | 10.21 (9.98 to 10.44) | 10.22 (9.97 to 10.47) |
| **Pearson r** | 0.56 (0.52 to 0.59) | 0.46 (0.43 to 0.48) | 0.41 (0.38 to 0.44) |
| **ICC** | 0.68 (0.64 to 0.71) | 0.60 (0.58 to 0.63) | 0.51 (0.47 to 0.54) |
| **LVEF ≤ 40%** |  |  |  |
| **Prevalence** | 538 (20.3%) | 1,052 (20.5%) | 763 (15.8%) |
| **AUROC (95%CI)** | 0.80 (0.77 to 0.82) | 0.76 (0.75 to 0.78) | 0.74 (0.72 to 0.75) |
| **AUPRC (95% CI)** | 0.50 (0.46 to 0.55) | 0.46 (0.43 to 0.49) | 0.39 (0.35 to 0.42) |
| **Sensitivity\*** | 0.66 (0.62 to 0.70) | 0.46 (0.43 to 0.49) | 0.48 (0.45 to 0.52) |
| **Specificity\*** | 0.82 (0.81 to 0.84) | 0.90 (0.89 to 0.91) | 0.89 (0.88 to 0.90) |
| **PPV\*** | 0.48 (0.45 to 0.52) | 0.54 (0.51 to 0.57) | 0.46 (0.43 to 0.49) |
| **NPV\*** | 0.90 (0.89 to 0.92) | 0.87 (0.86 to 0.88) | 0.90 (0.89 to 0.91) |
| **LVEF < 50%** |  |  |  |
| **Prevalence** | 829 (31.2%) | 1,643 (32.0%) | 1096 (22.7%) |
| **AUROC (95% CI)** | 0.77 (0.75 to 0.79) | 0.73 (0.72 to 0.74) | 0.71 (0.70 to 0.73) |
| **AUPRC (95% CI)** | 0.57 (0.54 to 0.61) | 0.55 (0.53 to 0.57) | 0.43 (0.41 to 0.46) |
| **Sensitivity†** | 0.57 (0.54 to 0.60) | 0.39 (0.36 to 0.41) | 0.42 (0.39 to 0.45) |
| **Specificity†** | 0.85 (0.84 to 0.87) | 0.92 (0.91 to 0.93) | 0.91 (0.90 to 0.91) |
| **PPV†** | 0.64 (0.60 to 0.67) | 0.69 (0.66 to 0.72) | 0.57 (0.53 to 0.60) |
| **NPV†** | 0.82 (0.80 to 0.83) | 0.76 (0.75 to 0.77) | 0.84 (0.83 to 0.85) |
| **Incident AFib at 5 years @ MHI (n = 3,121)** |  |  |  |
| **Prevalence** | 1,160 (37.2%) | - | - |
| **AUROC (95% CI)** | 0.61 (0.59 to 0.63) | - | - |
| **AUPRC (95% CI)** | 0.45 (0.42 to 0.48) | - | - |
| **PPV (95% CI) ‡** | 0.46 (0.43 to 0.48) | - | - |
| **NPV (95% CI) ‡** | 0.71 (0.68 to 0.73) | - | - |
| **Sensitivity (95% CI) ‡** | 0.59 (0.56 to 0.62) | - | - |
| **Specificity (95% CI)**<br>‡ | 0.59 (0.57 to 0.61) | - | - |
| **ACCO @ MHI (n = 1,021)** |  |  |  |
| **Prevalence** | 357 (35.0%) | - | - |
| **AUROC (95% CI)** | 0.85 (0.82 to 0.88) | - | - |
| **AUPRC (95% CI)** | 0.80 (0.77 to 0.84) | - | - |
| **PPV (95% CI) §** | 0.82 (0.78 to 0.87) | - | - |
| **NPV (95% CI) §** | 0.82 (0.80 to 0.85) | - | - |
| **Sensitivity (95% CI)**<br>§ | 0.63 (0.58 to 0.68) | - | - |
| **Specificity (95% CI)**<br>§ | 0.93 (0.91 to 0.95) | - | - |

Model outputs were conditioned on ECG signal content: across 996 QA pairs, LLM-as-a-judge scores fell from 0.40 with the correct ECG to 0.20 (zeroed) and 0.26 (a mismatched patient’s ECG). Outputs changed in over 91% of altered-input cases, and the model explicitly flagged signal absence in 243 of 996 zeroed examples versus 5 under normal input (Supplementary Tables 23 and 24).

Beyond automated scoring, we assessed the clinical quality of model-generated reports in a blinded reader study, in which board-certified cardiologists and cardiology trainees rated model and reference (cardiologist report) without knowing their source (Figure 1d,e). In the single-report study (Dataset A; 236 ECGs, 847 blinded ratings), the model matched the clinician reference. Mean aggregate quality on the 1-4 scale was 3.04 (95% CI 2.95 to 3.14) for the model versus 3.08 (95% CI 2.99 to 3.18) for the reference. Per axis, model versus reference was correctness 2.84 (2.73 to 2.95) vs 2.91 (2.79 to 3.02), completeness 3.04 (2.92 to 3.15) vs 3.12 (3.02 to 3.23) and clarity 3.26 (3.16 to 3.35) vs 3.22 (3.13 to 3.31). Both texts were flagged for a clinically significant error or hallucination at essentially the same rate (model 0.41, 95% CI 0.36 to 0.45; reference 0.39, 95% CI 0.35 to 0.44). This flag captured whether a reader judged the report to state a finding not supported by the tracing (a hallucination/false positive) or to omit a clinically important one (an error of omission); the two sources were closely matched on both failure modes - hallucinated or over-called findings in 0.30 of model versus 0.28 of reference ratings, and missed or mismeasured findings in 0.17 versus 0.18 - with most flagged issues, in both arms, judged to carry potential clinical harm. Paired head-to-head across the 230 ECGs rated on both sides was symmetric, with model win 0.32 (73/230; 95% CI 0.26 to 0.38), tie 0.35 (81/230; 95% CI 0.29 to 0.42) and reference win 0.33 (76/230; 95% CI 0.27 to 0.39), a net of −3 ECGs that was statistically indistinguishable from the reference (Wilcoxon p = 0.83, McNemar p = 0.87). Non-inferiority on correctness was directionally favorable (difference −0.03) but not formally established, as the lower confidence bound (−0.16) still crossed the pre-specified −0.05 margin at current coverage. Performance was category-dependent: the model was stronger on rhythm, infarct/ischemia, ACCO and culprit-artery question answering, the reference stronger on open interpretation, classification and conduction, and both were weakest on infarct/ischemia related question and answers (e.g. is there a Q wave on this ECG?).

The forced-choice study (Dataset B; 294 decoded case-level judgments) reproduced this equivalence: the model was preferred in 0.31 of cases (90/294; 95% CI 0.26 to 0.36), the reference in 0.28 (81/294; 95% CI 0.23 to 0.33) and the two were rated equivalent in 0.42 (123/294; 95% CI 0.36 to 0.48); among the 171 cases with a decided preference, the model was chosen in 0.53 (95% CI 0.45 to 0.60), again indistinguishable from chance (two-sided binomial p = 0.54). Reader reliability supported these read-outs. Within-reader test-retest on identical-content reports was high (mean |Δ| 0.21 on the aggregate score, 93% of re-ratings within ±1 point and 69% exact on all three axes) and between-reader agreement was moderate-to-substantial and axis-dependent (Gwet AC2, quadratic-weighted: clarity 0.67, completeness 0.53, correctness 0.41), the least reproducible judgment being the error/hallucination flag (Gwet AC1 0.28); cross-experience reader pairs disagreed more than same-experience pairs (mean |Δ| 0.76 vs 0.62). Representative case examples spanning ECG interpretation and rhythm classification, with paired model and reference reports and individual cardiologist ratings, are shown in Figure 4.

**Figure 4.**
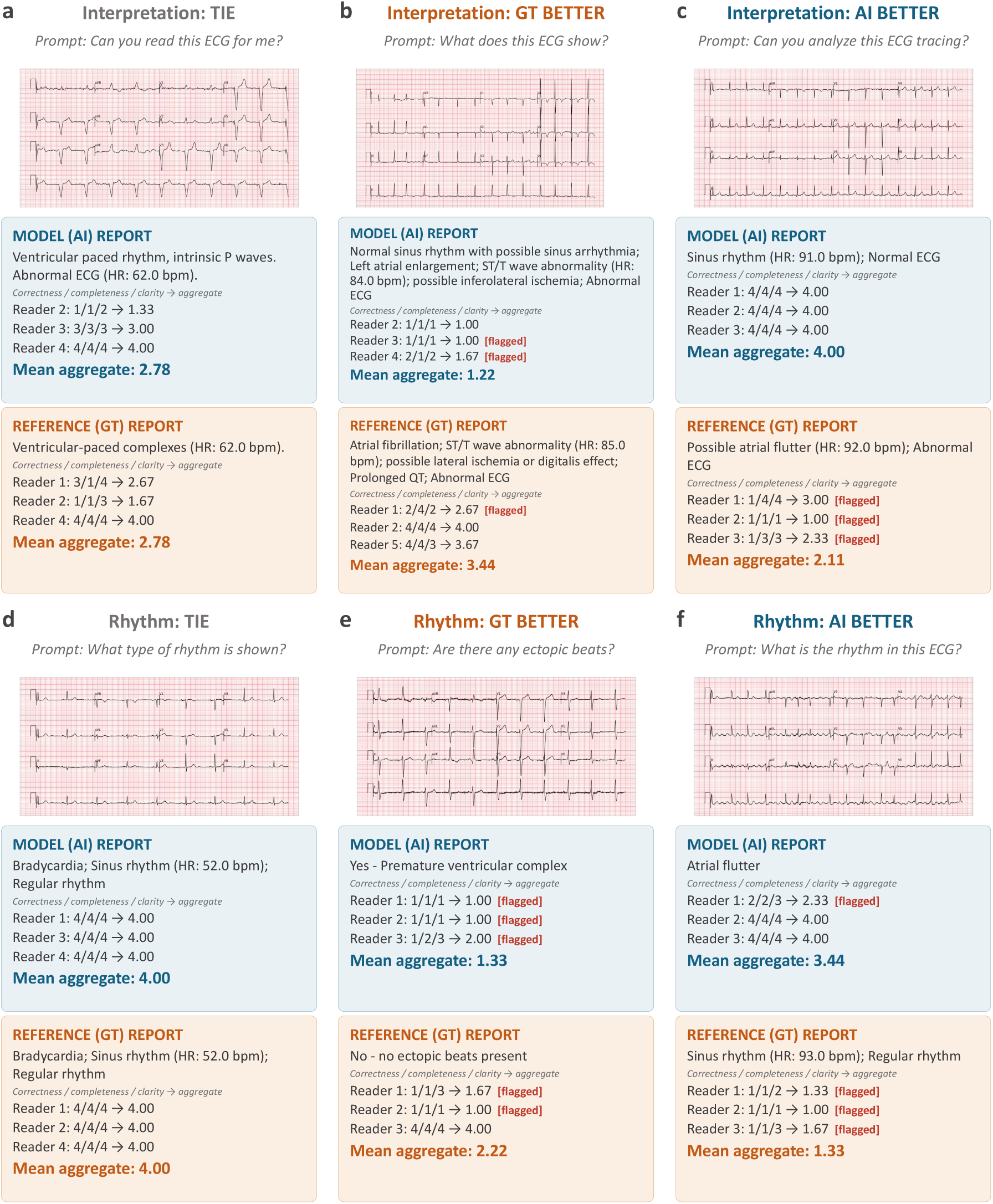
Representative examples of DeepECG-Tok reports versus reference reports with blinded cardiologist preferences. Six illustrative cases from the blinded reader study, arranged by task (rows) and adjudicated preference outcome (columns: tie, reference better, model better). Each panel shows the prompt posed to the model, the corresponding 12-lead ECG, and the paired free-text reports generated by DeepECG-Tok (MODEL (AI), blue) and the reference standard (REFERENCE (GT), orange); beneath each report, individual cardiologist ratings for correctness, completeness and clarity (1–4 scale) are listed with their mean aggregate, and “[flagged]” denotes a case a reader marked for review. a–c, ECG interpretation examples judged a tie (a), favoring the reference report (b), and favoring the model report (c). d–f, Rhythm classification examples judged a tie (d), favoring the reference (e), and favoring the model (f). Reports are shown verbatim; raters were blinded to report source at rating time.

### External Validation

In Stage 1, the frozen tokenizer generalized well: multi-label classification reached a macro-AUROC of 0.88 (95% CI, 0.87 to 0.91) on CLSA and 0.90 (0.89 to 0.91) on Harvard-Emory, versus 0.96 internally (Figure 2a; Supplementary Tables 16 and 25). Instruction-tuned interpretation also transferred, with weighted composite judge scores of 0.53 on CLSA and 0.50 on Harvard-Emory versus 0.71 internally; ischemia/infarction (0.69 and 0.66) and free-text interpretation (0.64 and 0.54) were the best-retained categories, while JSON interpretation attenuated most (0.42 and 0.38) (Figure 2c; Supplementary Table 26).External clinical-endpoint prediction was largely retained: LVEF estimation held at an MAE of approximately 10% (10.21% [95% CI, 9.98 to 10.44] on MIMIC-IV and 10.22% [9.97 to 10.47] on EchoNext) versus 8.41% (8.11 to 8.73) internally (Pearson r 0.46 [0.43 to 0.48] on MIMIC-IV and 0.41 [0.38 to 0.44] on EchoNext versus 0.56 [0.52 to 0.59] internally; Fisher r-to-z Z = 5.47 and 7.81, both p < 0.001), LVEF judge scores were comparable (0.76 [0.76 to 0.77] on MIMIC-IV and 0.77 [0.76 to 0.78] on EchoNext versus 0.82 [0.81 to 0.83] internally), and SHD detection on EchoNext-Mini (0.68 [0.67 to 0.70]) matched internal MHI (0.67 [0.65 to 0.69]) (Figures 2d and 3; Supplementary Table 22).

### Fairness Analyses

For Stage 1 classification in the pooled internal test set, ΔTPR was 0.01 (95% CI 0.01 to 0.01) and ΔFPR was 0.02 (95% CI 0.02 to 0.02) across sex, and ΔTPR was 0.024 (95% CI 0.022 to 0.025) and ΔFPR was 0.047 (95% CI 0.046 to 0.048) across age band. All values, including per-label maxima, fell below the pre-specified 0.10 tolerance in every cohort (Supplementary Table 27). Because equalized odds is prevalence-sensitive and base rates differ genuinely by age and sex, these estimates reflect descriptive parity rather than an absence of bias. For Stage 3 report generation, sex differences in the weighted composite judge score were consistent with parity: −0.01 (95% CI −0.06 to 0.04) pooled, 0.03 (95% CI 0.00 to 0.05) for MHI, and 0.00 (95% CI −0.05 to 0.06) for MIMIC-IV, with rank-biserial effect sizes at or below 0.04. Across age, the pooled comparison of patients above 75 versus below 55 years was null (−0.04, 95% CI −0.10 to 0.02), but averaged cohort effects in opposite directions: composite quality was lower in older MIMIC-IV patients (−0.08, 95% CI −0.14 to −0.02; rank-biserial 0.15), the only interval excluding zero, whereas the MHI difference was opposite in direction and indistinguishable from zero (0.03, 95% CI −0.01 to 0.07). All rank-biserial effect sizes were at or below 0.15 (Supplementary Table 28). Stage 3 subgroup analyses were exploratory, uncorrected for multiplicity, and not powered to exclude effects smaller than those observed.

## Discussion

DeepECG-Tok demonstrates that discrete tokenization of 12-lead ECG can support unified, clinically grounded interpretation, question answering, and clinical-endpoint prediction at scale, generalizing across four external cohorts (macro-averaged AUROC, CLSA 0.88; Harvard-Emory 0.90). Three contributions underpin this result: a novel adaptive RVQ tokenizer (QINCo) that yields a reusable discrete ECG representation, report generation that is state-of-the-art on aggregate natural-language-generation (NLG) metrics, evaluated under an ontology-grounded, cardiologist-validated LLM-as-a-judge that we release for the community with a dataset comprising ECGs and Q&As that can be used for benchmarking, and a single instruction-following model that performs multiple ECG tasks from one shared representation rather than a single model per task. The most direct evidence that these outputs are clinically usable comes from a blinded, fully-crossed reader study against a cardiologist reference. In single-report review, model interpretations were rated on par with cardiologist-adjudicated reports and answers, with a non-inferiority margin of −0.05 indicating parity. This extends a trajectory from cardiologist-level rhythm and abnormality detection ^10,11^ to free-text interpretation rated comparably to cardiologists’ reports and above legacy computerized reads ^36^. Whereas prior expert comparisons have evaluated classification accuracy or clinician performance with and without AI assistance, we evaluate the generated report itself as the unit of analysis, scored blind against the paired cardiologist report on the same scale under an imperfect-reference design.

The first contribution is a QINCo-based ECG tokenizer that produces reusable discrete ECG tokens serving every downstream task without a new representation model. Frozen embeddings evaluated on multi-label classification across 77 diagnostic conditions achieved a macro-averaged AUROC of 0.96 internally, outperforming the supervised DeepECG-SL and self-supervised DeepECG-SSL baselines and holding up across external cohorts without retraining. Because it optimizes for reconstruction rather than labels, the tokenizer learns morphology-preserving tokens that transfer across acquisition settings, so one frozen tokenizer serves interpretation, question answering, and endpoint prediction alike. QINCo also outperformed RVQ and vanilla VQ on reconstruction while reaching full codebook utilization, compared with 88.4% for RVQ and 38.9% for vanilla VQ. QINCo^17^ extends the neural-codec line of residual quantization like VQ-VAE^24^, SoundStream^25^, EnCodec^37^, DAC^38^. Adaptive per-stage codebooks keep codes active and avoid the collapse that caps VQ/RVQ utilization. This yields more reconstruction-faithful, morphology-preserving tokens than those produced by objective-mismatched schemes such as ECG-Byte^16^, whose frequency-driven byte-pair encoding is not optimized for waveform reconstruction or clinical representation.

The second contribution of DeepECG-Tok is state-of-the-art report generation and the framework that demonstrates it. The frozen tokenizer paired with a from-scratch GPT-2 decoder, and the same tokenizer conditioning a large language model, both outperformed the ECG-Byte^16^ baseline by a wide margin (Supplementary Table 10; macro-AUROC 0.70 for the tokenizer–GPT-2 decoder and 0.72 for the tokenizer-conditioned Llama 3.2-1B, versus 0.50 for ECG-Byte, with ROUGE, BLEU and METEOR substantially higher), and conventional natural-language metrics exceeded the previously reported ECG-GPT best by roughly 5 to 10 percent (Supplementary Table 29). On the ECG-GPT six-label protocol (Supplementary Table 21), DeepECG-Tok exceeded ECG-GPT on some labels and underperformed on others, most notably first-degree atrioventricular block (AUROC 0.66 versus 0.85); the improvement therefore holds on aggregate natural-language generation metrics and relative to ECG-Byte, and should not be interpreted as uniform superiority over ECG-GPT. These comparisons rest on an 847-term ECG ontology validated against board-certified cardiologists. Standard metrics such as BLEU and ROUGE measure surface-level lexical overlap and systematically penalize clinically correct synonyms (for example, “AFib with RVR” versus “atrial fibrillation with rapid ventricular response”). This mirrors the radiology literature, where lexical metrics correlate poorly with clinician judgment while LLM-based and entity-based metrics such as GREEN^39^, RadCliQ^40^ and MRScore^41^ correlate well, and our framework is the ECG analog. By combining deterministic rule-based scoring for structured outputs with semantic LLM adjudication for free-text narratives, it yields scores that better reflect clinical accuracy than textual similarity, and it is publicly available for other ECG and biosignal language models.

The third contribution dissolves the one-model-per-endpoint paradigm that has defined ECG deep learning: a single instruction-following model handles every task we evaluate, from narrative interpretation to continuous clinical endpoints, without a single task-specific head. Internal performance across 19 prompt categories follows a clinically interpretable gradient. Diagnostic categories with unambiguous morphological criteria scored highest on our LLM-as-a-judge benchmark, including chamber enlargement (0.86), rhythm disorders (0.81), and conduction disorders (0.73), while categories requiring clinical inference beyond the waveform scored lower, including AFib risk prediction (0.63) in patients in sinus rhythm, ACCO grading (0.59), and culprit artery identification (0.58). This hierarchy tracks differences in task observability from the ECG waveform and shows that the single model behaves coherently across task difficulty, although label quality and scoring format may also contribute. This unified formulation contrasts with two lines of prior work. Task-specific single-endpoint models achieve strong performance on individual endpoints, including left ventricular dysfunction^9,42^ and multi-site extensions^43^, structural heart disease^13^, and interpretation^4^ but each is narrow and not queryable in natural language. Other ECG-language models address only part of the task: MEIT^44^ generates reports only, ECG-Chat^45^ is conversational only, and PULSE^46^ and ECG-GPT^15^ operate on ECG images and can only do report generation but not questions and answering. None deliver both narrative interpretation and continuous clinical endpoints from one shared waveform representation, which is the capability DeepECG-Tok provides.

Across four independent external cohorts, endpoint-style, ECG-conditioned outputs transferred more robustly than open-ended report generation. Ischemia and infarction detection was the most robust interpretation category, with no significant internal-to-external drop, which is clinically encouraging because ST-segment changes and Q-wave patterns have well-defined waveform correlates. Attenuation was most pronounced for structured JSON interpretation, because exact field-level matching against a rigid schema is sensitive to differences in diagnostic label prevalence and reference-report formatting. The lower Harvard-Emory scores partly reflect a difference in reference standard, since the model’s training references at the MHI are cardiologist-authored whereas the Harvard-Emory reference reports are machine-generated by the Marquette 12SL algorithm (GE Healthcare), with residual differences in reporting conventions remaining even after stratified resampling.

Several limitations merit consideration. The linked clinical endpoint tasks, including AFib risk, SHD, ACCO, and culprit artery identification, were derived exclusively from the MHI cohort, where ECG data could be linked to echocardiographic and catheterization records; their generalizability to other institutions remains to be established through prospective multicenter validation. The retrospective study design and the absence of prospective deployment data mean that the operational impact of DeepECG-Tok in real clinical workflows has not yet been quantified.

The LLM-as-a-Judge framework, while validated against cardiologist adjudication, carries its own error rate and should itself be subject to ongoing calibration as new model generations become available. Report-derived labels also bound achievable Stage 1 performance, and because the imperfect-reference design scores the cardiologist reference on the same scale as the model, parity is parity against a fallible standard rather than against ground truth. The reader panel comprised six readers across two experience tiers, and because two of the three attending cardiologists were electrophysiologists, the senior tier skewed toward rhythm expertise, which is material given that the interpretation category was oversampled. Reader-study coverage was incomplete: Dataset A collected 847 of 1,416 target ratings (mean 1.8 of 3 per report across 472 reports) and Dataset B 294 of 711 (237 cases), so a substantial fraction of reports carry a single rating, which inflates measurement noise on the aggregate score and limits the Gwet AC1 and AC2 estimates that require double-rated items.

A signal-dependency ablation confirmed that outputs are conditioned on the ECG rather than text priors: zeroing or shuffling the input collapsed judge scores (0.40 to 0.20 and 0.26) and altered over 91% of outputs. Under zeroed input the model still returned a clinical answer in 75.6% of cases, flagging signal absence in 24.4% (243/996) and almost exclusively on open-ended prompts, while structured prompts consistently produced confabulated outputs. This silent-failure mode is a deployment concern: the structured-output pathways lack an abstention mechanism, so a deployed system would require an upstream signal-quality gate to withhold interpretation when the input is absent or corrupted, with reinforcement-learning fine-tuning to teach calibrated refusal as a complementary future direction, training on absent and mismatched (wrong-patient) ECGs so the model learns to abstain and explicitly state when the tracing does not support a finding. Token-level interpretability of QINCo codes is indirect, though this is mitigated by releasing the decoder so that any token sequence can be reconstructed and inspected.

Subgroup audits across sex and age indicate broadly comparable behavior, with mostly small differences, some of which nonetheless reach statistical significance: Stage 1 classification satisfies equalized odds, with all true-positive-rate and false-positive-rate disparities below the 0.10 tolerance we pre-specified as a descriptive screening bar rather than a validated criterion^3^, while Stage 3 report generation showed no meaningful sex difference and an age effect that differed by cohort: the pooled age comparison was null (−0.04, 95% CI −0.10 to 0.02), MIMIC-IV scored lower in patients above 75 years (−0.08, 95% CI −0.14 to −0.02; rank-biserial 0.15), and MHI ran in the opposite direction but was not distinguishable from zero (0.03, 95% CI −0.01 to 0.07). Because patients above 75 are the target population for the age-linked endpoints (incident AFib, reduced ejection fraction and structural heart disease), this reduction is clinically material regardless of mechanism; whether it reflects the greater case complexity of older patients or residual systematic bias cannot be resolved from the present analysis and warrants dedicated investigation. Subgroup analyses were limited to sex and age band; race and ethnicity were not evaluated because they are not collected at MHI, and a single-cohort fairness analysis was not reported. Fairness across paced rhythms, and across cohorts with participant-level demographics not available in the present external sources, has not yet been audited and is needed before clinical deployment.

A further limitation is that DeepECG-Tok does not yet match dedicated task-specific models on every endpoint. The unified instruction-following formulation trades a degree of per-task specificity for breadth, and on several categories the single model trails specialized single-endpoint baselines. Even so, it retains most of the performance of specialized single-endpoint models while spanning the full range of interpretation, question-answering, and endpoint prediction tasks from one shared representation. Narrowing this gap is a tractable direction for future work, through reinforcement learning with verifiable rewards such as Group Relative Policy Optimization (GRPO)^47^, which has recently demonstrated substantial gains in medical reasoning^48^ and multimodal clinical AI systems^49^, larger language-model backbones, alternative token-to-embedding projection architectures, and an expanded and more diverse ECG question corpus.

Built on reconstruction-optimized QINCo tokens, a single instruction-following model matches or approaches task-specific baselines across both morphological and endpoint tasks, and an ontology-grounded LLM-as-a-Judge framework provides a clinically meaningful evaluation standard where conventional lexical metrics fall short. External validation on four independent cohorts confirms that the approach generalizes beyond the training distribution, with endpoint-style outputs transferring most robustly. Prospective, multi-site validation of the MHI-derived clinical endpoints and adaptation of the judge to additional ECG acquisition systems are the essential next steps toward deployment.

## Author Contribution

R.B. and R.A. wrote most of the manuscript as well as conceived and conducted the experiments. R.A. and J.D. supervised the project conducted by R.B. S.S.S., A.S., A.N.-L., N.D., and O.T. provided critical review of the manuscript. J.B., B.M., N.D., G.M.-G., J.C.-T. and G.J. participated in the human validation study. R.B., J.D. and R.A. are the core contributors of this work; the order of the remaining authors (J.B., N.D., B.M., R.Ah., A.N.-L., G.J., S.S.S., G.M.-G., O.T., A.S. and J.C.-T.) was randomized. The opinions expressed in this manuscript are the authors’ own and do not necessarily reflect the views of the Canadian Longitudinal Study on Aging.

## Data Availability

The anonymized ECG dataset supporting the LLM-as-a-judge findings of this study will be made publicly available on PhysioNet (https://physionet.org/**) Upon Peer Review Publication.** The dataset comprises 25,000 twelve-lead ECGs selected from the DeepECG biobank at the Montreal Heart Institute (2006–2023), including raw signal files (NumPy .npy, 12 leads, 2,500 samples/lead at 250 Hz), standardized ECG images (PNG), and clinically validated question–answer pairs. Access requires PhysioNet credentialing, completion of CITI Data or Specimens Only Research training, and a signed data use agreement. Data are available from the Canadian Longitudinal Study on Aging (www.clsa-elcv.ca) for researchers who meet the criteria for access to de-identified CLSA data.

## Acknowledgements

This research was made possible using the data/biospecimens collected by the Canadian Longitudinal Study on Aging (CLSA). Funding for the Canadian Longitudinal Study on Aging (CLSA) is provided by the Government of Canada through the Canadian Institutes of Health Research (CIHR) under grant reference: LSA 94473 and the Canada Foundation for Innovation, as well as the following provinces, Newfoundland, Nova Scotia, Quebec, Ontario, Manitoba, Alberta, and British Columbia. This research has been conducted using Comprehensive Baseline and Follow-up 12-lead ECG data and Raw Data under Application ID 2401002 from the CLSA dataset: Comprehensive Follow-up 2 v2.0, Baseline, and Follow-up 1 ECG Images and Raw Data, under Application ID 2401002. The CLSA is led by Drs. Parminder Raina, Christina Wolfson and Susan Kirkland. The time and commitment of the participants to the CLSA study platform is gratefully acknowledged, without whom this research would not be possible.

## Code Availability

The complete evaluation framework including all judge implementations, the ECG medical ontology, scoring algorithms, and aggregation logic is available at https://github.com/HeartWise-AI/ECG_tokenizer/ and https://github.com/HeartWise-AI/ECG_LLM_Judge. The codebase is implemented in Python and requires the following dependencies: pandas, numpy, scipy, requests, and python-dotenv.

## Abbreviations

ACCO: Acute coronary occlusion
AFib: Atrial Fibrillation
AHA: American Heart Association
AI: Artificial Intelligence
AUPRC: Area under the precision recall curve
AUROC: Area under the receiver operating characteristic curve
AV: Atrioventricular
BERT: Bidirectional encoder representations from transformers
BLEU: Bilingual evaluation understudy
CI: Confidence interval
CLSA: Canadian Longitudinal Study on Aging
ECG: Electrocardiogram
ETC: ECG-text contrastive learning
ETG: ECG-conditioned text generation
ETM: ECG-text matching with hard-negative mining
FPR: False positive rate
HR: Heart rate
ICC: Intraclass correlation coefficient
JSON: JavaScript object notation
LBBB: Left bundle branch block
LLM: Large Language Model
LoRA: Low-rank adaptation
LVEF: Left ventricular ejection fraction
MAE: Mean Absolute Error
METEOR: Metric for evaluation of translation with explicit ordering
MHI: Montreal Heart Institute
MI: Myocardial infarction
MIMIC: Medical Information Mart for Intensive Care
NPV: Negative predictive value
PPV: Positive predictive value
QA: Question-Answer
Q-Former: Query transformer
QINCo: Quantization with Implicit Neural Codebooks
RBBB: Right bundle branch block
ROUGE: Recall-oriented understudy for gisting evaluation
RVQ: Residual Vector Quantization
SHD: Structural heart disease
TPR: True positive rate
TTE: Transthoracic echocardiography
VQ: Vector Quantization

