## Supplementary Materials for "A unified 12-lead ECG-language model for interpretation and clinical-endpoint prediction"

### Supplemental Methods

#### S1. ECG preprocessing

All input formats (XML, WFDB, HDF5, DICOM) were parsed to yield a NumPy array of shape (12, 2500) representing the 12 leads at 250 Hz in standard order (I, II, III, aVR, aVL, aVF, V1–V6). Processing proceeded in the following sequential steps:

***Resampling.*** Recordings acquired at 500 Hz were downsampled to 250 Hz using a polyphase anti-aliasing filter.

***Baseline wander correction.*** Signals were transformed to the frequency domain via FFT. A zero-phase 1 Hz high-pass Butterworth filter was applied conditionally: only when spectral energy below 1 Hz exceeded that of the 1–30 Hz band by more than 10-fold, preserving signals with stable baselines.

***Amplitude normalization.*** Mean spectral power in the 1–30 Hz band was compared against a reference curve to derive a per-recording scaling factor (factor = target power / mean spectral power), and all frequency components were scaled accordingly before IFFT reconstruction.

***Powerline artifact removal.*** Spectral peaks at 50 Hz, 60 Hz, and their harmonics (integer multiples within 5% tolerance) were identified using a sliding-window threshold (mean + k × SD). Detected peaks were flattened to a local baseline estimated by LOESS smoothing. The cleaned signal was reconstructed via IFFT.

#### S2. BERT report labeler

***Rationale and ontology.*** We follow the previously published model and methods[3]. Free-text ECG reports at MHI (French and English) and MIMIC-IV (English)[1] are too variable in phrasing, abbreviation, and negation for manual annotation at foundation-model scale. We therefore used a BERT-based classifier to map diagnostic statements to a 77-condition ontology organized into six AHA-aligned categories[2] (Rhythm Disorders, Conduction Disorders, Chamber Enlargement or Hypertrophy, Ischemia and Infarction, Pericarditis, Other Diagnoses), following our previously published DeepECG framework (Supplementary Table 8).

***Annotation and training.*** Two cardiologists independently annotated 7,400 diagnostic statements (MHI n = 4,200; MIMIC-IV n = 3,200), stratified to include rare conditions, with a third cardiologist adjudicating discrepancies (Cohen's kappa > 0.80 across all categories). A pretrained BERT-base encoder[3] (multilingual variant for French MHI statements) was fine-tuned for multi-label classification with one sigmoid output per condition, using binary cross-entropy loss, AdamW optimization[4] and early stopping on a held-out validation split.

***Performance and label propagation.*** The classifier achieved AUROC > 0.98 across all 77 conditions (minimum 0.98, U wave; median > 0.99) and was applied to the full MHI and MIMIC-IV datasets to generate structured multi-label annotations. Conditions semantically subsumed elsewhere (e.g., RAVL ≥ 11 mm under left ventricular hypertrophy) were removed from "Other Diagnoses" to avoid redundancy. The resulting labels served as ground truth for all downstream probing, tokenizer evaluation, and instruction-tuning analyses.

### S3. ECG tokenizer architecture and training

#### Architecture

The encoder consisted of three 1-D convolutional blocks with residual connections[5]. The first block mapped 12 input ECG leads to 32 channels using a convolutional layer with kernel size 4 and stride 2, followed by max pooling with kernel size 2 and stride 2 and a GELU[6] activation. The second block expanded to 64 filters with the same pattern. The third block expanded to 128 channels using a convolutional layer with kernel size 4 and stride 2, without pooling. Each block included a residual refinement sub-block composed of two Conv1d layers with kernel size 3 and BatchNorm[7]. This architecture reduced each 10-second, 2,500-sample input ECG sequence to a latent sequence of length 82 with 128 channels, corresponding to approximately 30-fold temporal down-sampling.

The RVQ bottleneck applied 8 sequential codebooks[8], Q1-Q8, each with 512 entries, to the latent sequence. Unlike standard RVQ, QINCo[9] conditioned each codebook on the evolving partial reconstruction via a small implicit neural network (MLP), enabling finer morphological discretization at equivalent codebook capacity. The decoder mirrored the encoder with three transposed convolutional layers (128 to 64 to 32 to 12 channels) interleaved with GELU activations and nearest-neighbor 2x upsampling to reconstruct the original 12-lead waveform. The complete tokenizer (encoder, QINCo residual quantizer, and decoder) contained approximately 0.9 million parameters.

#### Training objective

The tokenizer was trained using a composite objective combining mean absolute reconstruction error across all leads and time points with a vector-quantization[10] commitment penalty that encouraged encoder outputs to remain close to their assigned codebook entries. The commitment term used a weight of 0.25.

#### Training configuration

The tokenizer was trained for 10 epochs on 1,914,615 ECGs from MHI, MIMIC-IV and CODE-15[11] using AdamW with a lr = 3e-4 and weight decay = 1e-4. We used a cosine warm restart schedule[12] and linear warmup over the first 10% of training (approximately 14,957 steps). Batch size was 32 per GPU. Training was conducted on 4 x NVIDIA H200 GPUs over approximately 18 hours.

#### S4. Tokenizer-to-report generation

As auxiliary validation that the learned discrete ECG representation could support diagnostic language generation, we trained autoregressive text decoders conditioned on the frozen quantized ECG sequence. The ECG tokenizer, including the convolutional encoder and QINCo residual quantizer, was kept frozen throughout these experiments so that performance reflected the information retained in the learned token sequence rather than further adaptation of the signal encoder.

Each ECG was passed through the frozen QINCo tokenizer to obtain the quantized latent of shape (128 channels, 82 time steps), as defined in S3. A lightweight SequenceAdapter then mapped this representation to a single conditioning vector in the decoder embedding space. Each of the 128 channel positions, represented by its 82-dimensional code vector across time, was first projected to an intermediate dimension d_int (d_int = 384 for the GPT-2[13] decoder, d_int = 1,024 for the Llama 3.2-1B[14] decoder) using a position-wise feed-forward block (linear, RMSNorm, GELU, dropout 0.2). A learnable positional embedding of shape (128, d_int), initialised from a normal distribution with standard deviation 0.02, was added across the channel axis. The 128-position sequence was then contextualised using a single multi-head self-attention layer (8 heads, embedding dimension d_int), followed by a residual connection and RMSNorm. The 128 contextualised tokens were aggregated by mean pooling to produce a single d_int-dimensional vector. A final feed-forward block projected this vector from d_int to the decoder embedding dimension d_dec (d_dec = 768 for GPT-2; d_dec = 2,048 for Llama 3.2-1B), again followed by RMSNorm, GELU, and dropout 0.2, and scaled by a learnable scalar.

We evaluated two pretrained autoregressive decoders, GPT-2 and Llama 3.2-1B. For each decoder, the projected ECG vector replaced the embedding row of a special <ecg> token in the language-model input, providing ECG conditioning as a single prefix embedding before the report tokens. The decoder was trained to predict report tokens autoregressively, conditioned on the ECG prefix and all preceding report tokens. The base decoder weights were kept frozen, and only the SequenceAdapter and decoder LoRA[15] parameters were optimized.

#### S5. Q-Former ECG-language alignment

Stage 2 trained a Q-Former[16] bridge (198.7M parameters) to align frozen discrete ECG tokens with the MedGemma 4B-IT[17] text representation space with both the ECG tokenizer and the language backbone fully frozen; the bridge cross-attended to the frozen ECG encoder every two layers, and text tokens never attended directly to ECG embeddings. Query outputs were pooled by RMSNorm followed by a sigmoid gate and routed through separate ECG-side and text-side projection heads for the contrastive and matching objectives, while a dedicated text embedding layer (201.3M parameters, tied with the LM head, initialized from MedGemma's embeddings but trained independently) produced text representations via masked mean pooling.

Three objectives were optimized jointly. ECG-text contrastive learning (ETC) used a SigLIP[18]-style symmetric binary cross-entropy with a learnable temperature, with boosted alpha weights on the 50 rarest text IDs to prevent collapse toward majority classes and a pre-encoded text embedding bank of 2,048 entries refreshed every 256 steps that contributed 5 additional negatives per ECG. ECG-text matching (ETM) trained a 2-class linear head on fused query-text representations, with hard negatives drawn from dynamic in-batch mining (top 3 most similar non-matching texts per ECG), 12 clinically curated families covering known waveform confusability across 5 diagnostic groups (e.g., ST elevation versus depression in the same lead territory, VT versus SVT with aberrancy, varying AV block degrees), and yes/no answer flips for binary diagnostic questions. ECG-conditioned generation (ETG) used teacher-forced next-token prediction conditioned on the query-token outputs. ETM weight was linearly ramped from 0 to 1.0 over the first 2,000 steps and ETG was held at zero until step 2,000 then ramped to 1.0 by step 4,000, letting contrastive representations stabilize before harder objectives were engaged. Training used 761,386 MHI ECGs paired with 222 text entries (74 atomic diagnostic statements plus 148 binary QA pairs) yielding 9.57M ECG-text pairs (a mean of 12.6 text targets per ECG), with validation on 94,948 pairs from 7,565 held-out ECGs; optimization used AdamW (learning rate $1.4 \times 10^{-4}$, weight decay 0.01, batch size 120, bfloat16) for 10 epochs.

Twelve hyperparameters were explored via Bayesian sweep (Weights & Biases) with Hyperband early stopping (min_iter = 3, $\eta = 2$) across the ranges shown in Supplementary Table 6. The best configuration was selected on validation contrastive loss and used to initialize the instruction-aware Q-Former in Stage 3.

### S6. Instruction-tuned MedGemma model

#### Overview

Stage 3 combined the frozen ECG tokenizer and the Stage 2-initialized Q-Former bridge with MedGemma 4B-IT for multi-task ECG instruction tuning. The Q-Former was warm-started from the best Stage 2 checkpoint and remained trainable during instruction tuning. The MedGemma 4B-IT backbone was kept frozen except for LoRA modules inserted into the top 12 of its 34 transformer layers. The ECG encoder, QINCo residual quantizer, MedGemma token embeddings and all transformer layers below the LoRA-adapted window remained frozen throughout training. The Stage-3 bridge used 32 query tokens and 12 attention heads (expanded from the 16 tokens and 8 heads used in Stage 2), and all eight codebooks were retained rather than a reduced subset, preserving the fine morphological detail required by interval-estimation and LVEF tasks.

#### Trainable parameters

LoRA modules were applied to the query, key, value and output attention projection matrices of each adapted MedGemma layer, with rank $r = 32$, scaling factor $\alpha = 64$ and dropout 0.05. Given MedGemma 4B-IT's hidden size of 2,560 and grouped-query attention configuration, each adapted layer contributed 524,288 LoRA parameters, yielding 6,291,456 trainable LoRA parameters across the 12 adapted layers. The instruction-aware Q-Former contributed an additional 200,079,881 trainable parameters. The final Stage 3 model therefore contained 206.4M trainable parameters, corresponding to approximately 4.8% of the 4.3B-parameter combined model.

#### Prompt formatting

Each training example was rendered into a three-turn Gemma chat using literal turn delimiters, with BOS and EOS added by its tokenizer:

**<start_of_turn> system**
You are an expert cardiologist. You interpret ECGs and answer in a concise, structured way.
**<end_of_turn>**
**<start_of_turn> user**
**<start_of_image>**
Question: {question}
Respond concisely with the key finding or answer.
**<end_of_turn>**
**<start_of_turn> model**
{answer}
**<end_of_turn>**

The system message was held constant across all instruction-tuning samples. The user turn began with a single <start_of_image> sentinel token, followed by a blank line, the natural-language question drawn from the prompt column of the training corpus, and a fixed response-format suffix. The model turn contained the ground-truth answer at training time and was empty at inference and generated autoregressively. All examples in the canonical training corpus were free-text question-answer pairs spanning ECG interpretation, structured diagnostic outputs, rhythm classification, and clinical endpoint prediction.

#### ECG soft-prompt insertion

The 32 ECG query-token embeddings emitted by the Q-Former, already projected into MedGemma 4B-IT's 2,560-dimensional input space, were spliced into the embedded token sequence immediately after the position of the <start_of_image> token. The 32 ECG query-token embeddings were inserted directly into the input embedding sequence (after the embedding lookup of the text tokens) at the position immediately following the <start_of_image> token. No discrete placeholder token was reserved for the ECG positions; the language model received the fused embedding sequence via the inputs_embeds interface, with input_ids, attention_mask, and labels padded to the same length for tensor-shape alignment. Each fused input sequence therefore comprised the 32 ECG query-token embeddings plus the chat-template text tokens, giving a median of 110 tokens (range 87–316; maximum context 672) passed to DeepECG-Tok per example.

#### Loss masking

The autoregressive next-token-prediction objective was applied only to tokens belonging to the model turn. Every position from the start of the sequence through the <start_of_turn>model\n boundary, the 32 ECG soft-prompt positions, and any right-side padding were assigned the ignore index (label = −100); only the answer tokens and the closing <end_of_turn> of the model turn contributed to the cross-entropy gradient.

#### Training corpus

Training used 7,274,356 ECG-question-answer pairs from MHI and MIMIC-IV, supplied through a single parquet file with a per-example sample_weight column read by a WeightedDistributedSampler to upsample minority diagnostic categories. Validation used a held-out 49,776-pair corpus drawn from the same two sources; mid-epoch validation was performed every 1,000 optimizer steps on a 2,000-pair subset balanced across prompt categories. The training and validation corpora were constructed from the Stage 1 patient partitions, with no patient overlap between splits.

#### Optimization

Training used AdamW with separate parameter groups for the LoRA adapters and the Q-Former bridge. The LoRA group used learning rate 5e-5 and weight decay 1e-5; the Q-Former group used learning rate 5e-4 and weight decay 1e-5. A cosine schedule with linear warmup over 25.6% of total steps was applied to both groups. Per-GPU batch size was 16 with gradient accumulation of 16 steps over 4 NVIDIA H200 GPUs in distributed data-parallel mode, yielding an effective global batch size of 1,024. The maximum input sequence length was 768 tokens (640 prompt and 128 generation). Training ran for 1 epoch.

#### Hyperparameter selection

Sixteen hyperparameters were explored via Bayesian sweep with Hyperband early stopping and progressive resource allocation across trials, across the ranges shown in Supplementary Table 7. The best configuration was selected on an LLM-as-a-judge composite score on the internal validation set and used to produce the instruction-tuned model evaluated in the main text.

### S7. LLM-as-a-judge framework

Standard natural language generation metrics (BLEU[19], ROUGE[20], METEOR[21]) measure lexical overlap and systematically penalize clinically correct predictions that use synonymous terminology. "Atrial fibrillation with rapid ventricular response" and "AFib with RVR" are clinically identical yet score near-zero by lexical similarity. We therefore developed a hierarchical evaluation framework combining deterministic rule-based scoring for structured outputs with LLM-adjudicated semantic evaluation[22] for free-text narratives.

##### Routing Architecture

All predictions are routed to one of two evaluation pathways based on prompt category metadata (Supplementary Table 30). Structured and categorical outputs (JSON fields, binary labels, numeric measurements) undergo deterministic field-by-field comparison. Free-text narrative interpretations are routed to the LLM adjudicator for semantic evaluation with ontology-assisted synonym resolution.

##### Deterministic Evaluation

Categorical fields with a single value are scored by exact match after ontology-based normalization. Multi-value categorical fields (e.g., a list of rhythm findings) are scored by Jaccard similarity:

score = |prediction ∩ ground_truth| / |prediction ∪ ground_truth|

Numeric predictions are evaluated against clinically derived tolerance thresholds (Supplementary Table 13). Predictions within tolerance receive full credit; predictions outside tolerance receive partial credit proportional to deviation magnitude:

score = max(0, 1 − |prediction − ground_truth| / (tolerance × 2))

Predictions exceeding twice the tolerance threshold receive zero credit.

**LLM Adjudicator**

Free-text outputs were evaluated by MiniMax-M2.5[23] accessed via the Fireworks AI inference API at temperature 0.0 to ensure deterministic reproducibility with structured JSON format. The model was prompted to act as a board-certified cardiologist assessing whether a model-generated ECG interpretation accurately captures findings from a reference cardiologist report. For each example, the adjudicator received the ground-truth interpretation, the model prediction, and a relevant excerpt from the ECG ontology. It enumerated distinct clinical findings in each, resolved synonyms via ontology lookup, and classified all findings as: identified (semantically present in both), missed (in ground truth only), or hallucinated (in prediction without ground-truth support). Output was returned as structured JSON.

**ECG Medical Ontology**

To enable accurate synonym resolution, we constructed an ECG terminology ontology comprising 847 validated term mappings across 12 diagnostic categories. The ontology additionally encodes parent-child relationships that enable partial credit for hierarchically related terms: intraventricular conduction delay mapping to LBBB (0.7 credit), supraventricular tachycardia to atrial fibrillation (0.5 credit), and ischemic changes to anterior STEMI (0.6 credit). Representative synonym clusters include: atrial fibrillation (AFib, AF, A-fib, auricular fibrillation, fibrillating atria); LBBB (left bundle branch block, left BBB, L-BBB, complete LBBB, CLBBB); STEMI (ST-elevation MI, ST elevation myocardial infarction, acute STEMI, STE-ACS).

**Score Computation**

The interpretation score treats omission and commission errors symmetrically:

score = |identified| / (|identified| + |missed| + |hallucinations|)

For continuous predictions (heart rate, LVEF), performance is additionally summarized by MAE, Pearson r, Spearman rho, and ICC(2,1) for absolute agreement. ICC thresholds follow standard conventions: >0.75 excellent, 0.50-0.75 moderate, <0.50 poor. Final composite scores are computed as weighted averages across all prompt categories, with weights reflecting clinical importance (Supplementary Table 30). ACS severity tasks use a six-class hierarchical schema (No Disease, Obstructive No Acute, Chronic Occlusion, Acute Incomplete, Acute Complete, Acute Other); per-class sensitivity and specificity are reported to characterize systematic misclassification patterns.

Weights were picked arbitrarily due to task complexity with interpretation being weighted higher than other categorical or regression answers.

### S8. Signal dependency analysis

To assess whether model outputs were conditioned on ECG signal content rather than text priors, we designed a controlled perturbation experiment on the MHI held-out test set. From 259,233 test ECGs, 1,669 were sampled with stratification across all 77 diagnostic labels. Standard QA generation produced 6,661 question-answer pairs, from which 996 were retained after stratified subsampling by prompt category (interpretation: 243, classification: 243, JSON interpretation: 243, rhythm: 133, ischemia/infarction: 110, other: 14, conduction: 10).

Each QA pair was evaluated under three input conditions using identical prompts, model weights, and inference parameters (MedGemma 4B-IT with LoRA, 8-codebook QINCo tokenizer, NVIDIA H200 GPU):

1. Normal: the correctly preprocessed, power spectrum analysis (PSA)-normalized ECG waveform.
2. Zeroed: an all-zeros tensor of identical shape (12 channels, 2,500 samples), replacing the ECG input entirely. This condition tests whether the model generates clinically plausible outputs in the absence of any signal, which would indicate reliance on text-prompt priors rather than ECG features.
3. Shuffled: a randomly selected ECG from a different patient in the test set, paired with the original patient's prompt. This condition tests whether the model produces outputs specific to the input waveform or generates generic responses independent of signal content.

Model outputs were evaluated across four complementary axes. First, clinical accuracy was scored using the LLM-as-a-judge framework (overall and per-category). Second, lexical similarity to ground truth was computed using ROUGE-1, ROUGE-2, ROUGE-L, BLEU-1, BLEU-4, and METEOR. Third, cross-condition output divergence was quantified by exact-match rate and pairwise ROUGE-L similarity between outputs generated under different conditions for the same prompt. Fourth, signal-absence detection was assessed by counting outputs containing explicit refusal statements (e.g., "analysis impossible," "no QRS detectable," "cannot analyze") and measuring mean output length in characters.

### S9. Baselines

#### DeepECG-SL and DeepECG-SSL

DeepECG-SL[24] and DeepECG-SSL[24] were used as supervised and self-supervised baselines for evaluating representation quality. DeepECG-SL is an end-to-end EfficientNetV2[25] classifier trained on 12-lead ECGs, whereas DeepECG-SSL is a transformer pre-trained on raw ECG signals and fine-tuned for the same 77-condition classification task. Both models were evaluated using released checkpoints without retraining.

All models were evaluated on the same patient-disjoint internal test set (416,467 ECGs from MHI and MIMIC-IV) using identical preprocessing and input manifests. Ground-truth labels were derived using the validated 77-condition BERT-based classifier applied to cardiologist reports, with published per-class thresholds applied uniformly across all models.

Performance was assessed using per-class AUROC and AUPRC, with macro-averaged values reported across six diagnostic categories and the full 77-condition label set. Confidence intervals were estimated using the same bootstrap protocol as for DeepECG-Tok (1,000 iterations; seed = 42). No model retraining, threshold adjustment, or evaluation modification was performed, ensuring a direct comparison of representation quality across models.

#### ECG-Byte

For evaluating tokenization-based ECG-to-text generation, we compared against ECG-Byte[26] using its discrete signal-token representation framework. To enable a controlled comparison, we matched the LLM backbone and parameter-efficient fine-tuning setup across methods. Specifically, both ECG-Byte and our tokenizer-based approach were paired with a Llama 3.2-1B decoder and trained using LoRA adapters applied to the query and value projection matrices with rank 16.

The two approaches differ only in how ECG information is provided to the language model. ECG-Byte represents each waveform as a sequence of discrete signal tokens inserted into the text vocabulary, whereas our approach first discretizes the ECG using a learned vector-quantized tokenizer (QINCo), then maps the resulting token sequence to a continuous embedding via a sequence adapter for conditioning the language model. This design isolates the effect of the conditioning mechanism from the language model and fine-tuning budget.

Training was performed on 1.49 million ECG–report pairs from the interpretation subset of the internal dataset for one epoch, using identical preprocessing, prompt formatting, and patient-disjoint train–test splits. Evaluation was conducted on a held-out set of 10,000 ECGs using ROUGE-1, ROUGE-L, BLEU-4, and METEOR. In addition, generated reports were mapped to the 77-condition ontology using the validated BERT-based classifier, and diagnostic agreement with reference labels was summarized using macro-averaged AUROC and AUPRC. Confidence intervals were computed using the same bootstrap protocol described in the Statistical Analysis section.

#### ECG-GPT

ECG-GPT[27] was used as the primary published ECG report-generation baseline because it generates free-text ECG interpretations and reports structured label-assessment metrics on PTB-XL[28], a public dataset that supports direct comparison. We evaluated our models under ECG-GPT's published structured label-assessment protocol, using PTB-XL v1.0.3 as the shared test set and the six rhythm and conduction labels reported in ECG-GPT Table 4: atrial fibrillation, sinus tachycardia, sinus bradycardia, complete left bundle branch block, complete right bundle branch block and first-degree atrioventricular block.

For our models, each PTB-XL ECG was passed through the DeepECG-Tok tokenizer and decoded into one free-text interpretation using either a GPT-2 decoder or MedGemma 4B-IT. GPT-2 was evaluated on all 21,799 PTB-XL ECGs, whereas MedGemma was evaluated on the 10,454 ECGs retained by the question-answer preprocessing pipeline and restricted to the interpretation prompt category. Generated reports were converted into binary label predictions using ECG-GPT's published keyword vocabulary and negation-handling rules. Ground-truth labels were derived from PTB-XL SCP codes, with mappings chosen to match ECG-GPT's reported label prevalences.

Performance was summarized using the same metrics reported by ECG-GPT, including AUROC and AUPRC with 95% confidence intervals. Because ECG-GPT's keyword-extraction protocol produces binary predictions, the reported AUROC corresponds to balanced accuracy, defined as the average of sensitivity and specificity; we preserved this convention to maintain direct comparability with ECG-GPT Table 4. Confidence intervals for our models were estimated using 1,000 ECG-level bootstrap resamples with seed 42. ECG-GPT comparator values were taken directly from the published Table 4 and were not reimplemented or re-estimated.

### Supplemental Tables

#### Supplementary Table 1 | Patient-level data splits and stage-by-stage data usage across the DeepECG-Tok pipeline

##### Supplementary Table 1a | Stage 1: ECG tokenizer

| Cohort | Training ECGs | Training Patients | Test ECGs | Test Patients |
|---|---|---|---|---|
| MHI | 1,017,719 | 184,210 | 259,233 | 51,213 |
| MIMIC-IV | 551,099 | 112,539 | 157,234 | 31,902 |
| Code-15 | 345,797 | 233,770 | - | - |

| | | | | |
|---|---|---|---|---|
| Total | 1,914,615 | 530,519 | 416,467 | 83,115 |

Supplementary Table 1b | Q-Former pre-training

| **Split** | **Cohort** | **ECG-text pairs** | **ECGs** | **Patients** |
|---|---|---|---|---|
| Training | MHI | 9,565,410 | 761,386 | 136,676 |
| Validation | MHI | 94,948 | 7,565 | 4,963 |
| Test | MHI | 2,698,026 | 215,345 | 38,811 |

Supplementary Table 1c | LLM fine-tuning data splits

| **Cohort** | **Training QA pairs** | **Training ECGs** | **Training patients** | **Test QA pairs** | **Test ECGs** | **Test patients** |
|---|---|---|---|---|---|---|
| MHI | 4,744,609 | 664,697 | 133,116 | 24,995 | 5,000 | 3,858 |
| MIMIC-IV | 2,529,747 | 423,204 | 85,203 | 24,781 | 5,000 | 3,585 |
| Total | 7,274,356 | 1,087,901 | 218,319 | 49,776 | 10,000 | 7,443 |

### Supplementary Table 2 | Multi-modal clinical tasks (MHI cohort only)

| **Task** | **N** | **Classes** | **Prevalence** | **Weight** | **Example Prompt → Answer** |
|---|---|---|---|---|---|
| afib_risk | 499,839 | High (AFib within 2 years) | 28.2% | 2.0 | “Will this patient develop AFib?” → “Low risk” |
| | | Moderate (AFib within 5 years) | 3.2% | 2.0 | |
| | | Low | 68.6% | 1.0 | |
| shd | 254,460 | Present | 41.4% | 1.5 | “Does this patient have |

| | | | | | |
|---|---|---|---|---|---|
| | | Absent | 58.6% | 1.0 | structural heart disease?” → “No SHD detected” |
| lvef | 197,422 | Severely reduced | 7.2% | 4.0 | “What is the LV function?” → “LVEF 28% (severely reduced)” |
| | | Moderately reduced | 14.8% | 3.0 | |
| | | Mildly reduced | 17.4% | 2.0 | |
| | | Normal | 60.6% | 1.0 | |
| acs_severity | 28,373 | No disease | 14.8% | 1.0 | “Is there acute coronary occlusion?” → “No obstructive CAD without acute occlusion” |
| | | Obstructive (no acute) | 65.0% | 1.0 | |
| | | Chronic occlusion | 14.1% | 2.0 | |
| | | Acute complete | 2.9% | 3.0 | |
| | | Acute incomplete | 3.0% | 3.0 | |
| | | Acute other (culprit not documente d) | 0.1% | 3.0 | |
| culprit_artery | 919 | LCX | 12% | 4.0 | “What is the culprit artery?” → “Proximal LAD with complete occlusion” |
| | | LAD | 31% | 1.5 | |
| | | RCA | 57% | 1.0 | |

### Supplementary Table 3 | Standard ECG interpretation tasks

| **Task** | **N** | **Type** | **Example Prompt** | **Example Answer** |
|---|---|---|---|---|
| interpretation | 1,491,307 | Free text | “What does this ECG show?” | “Atrial fibrillation (HR: 75 bpm); Right bundle branch block” |
| json_interpretation | 1,422,249 | JSON | “Output JSON ONLY with keys: RHYTHM, CONDUCTION...” | {“RHYTHM”:[“A Fib”], “CONDUCTION”:[“RBBB”], “heart_rate_bpm”:75} |
| classification | 1,394,209 | Binary | “Is this ECG normal or abnormal?” | “No – Abnormal ECG; Pathological findings: Atrial Fibrillation” |
| category_rhythm | 1,042,305 | Single-label | “What is the rhythm in this ECG?” | “Irregularly irregular rhythm (HR: 75 bpm); Atrial Fibrillation” |
| category_conduction | 285,167 | Multi-label | “What type of block is present if any” | “Right bundle branch block” |
| category_infarct_ischemia | 425,023 | Multi-label | “Is there any ST elevation or depression?” | “Yes – ST depression (lateral leads); Q waves in inferior leads” |
| category_chamber_enlargement | 42,296 | Multi-label | “Does this show ventricular hypertrophy?” | “Yes – LVH” |
| category_pericarditis | 1,590 | Binary | “Is there diffuse ST elevation” | “Yes – pericarditis with ST changes” |

| | | | | |
|---|---|---|---|---|
| category_other | 184,398 | Multi-label | “Is there anything else abnormal?” | “Yes – T wave inversion (septal V1-V2); T wave inversion (anterior V3-V4)” |
| ecg_interval | 1,516 | Integer | “What is the patient’s heart rate?” | “60 bpm” |
| random_finding_question | 1,234 | Binary | Is there 1st degree AV block? | “Yes” |
| localization_qrs_axis | 1,152 | Categorical | “Is there axis deviation?” | “Yes – Left axis deviation (-30° to -90°)” |
| localization_q_wave | 329 | Localized | “Are there Q waves suggesting old infarction?” | “Yes – Q wave in septal leads V1, V2; anterior leads V3-V4” |
| localization_st_depression | 18 | Localized | “Are there ST depressions in any leads?” | “Yes – ST depression in lateral leads I, aVL, V5-V6” |
| localization_t_wave | 489 | Localized | “Can you identify T wave changes?” | “Yes – T wave inversion in lateral leads I, aVL, V5, V6” |
| urgency_assessment | 70 | Triage | “Is this an emergency ECG finding?” | “URGENT: Immediate intervention required – ST elevation (anterior V3-V4)” |

Supplementary Table 4 | Comparison of vector quantization strategies for ECG tokenization

| | VQ | RVQ | QINCo |
|---|---|---|---|
| **Codebook Size** | 512 | 512 | 512 |
| **Number of Quantizers** | 1 | 8 | 8 |
| **Loss Function** | MAE + commitment loss | MAE + commitment loss | MAE + commitment loss |
| **Optimizer** | AdamW | AdamW | AdamW |
| **Learning rate** | 3e-4 | 3e-4 | 3e-4 |
| **Batch size** | 32 | 32 | 32 |
| **Epochs** | 10 | 10 | 10 |
| **Scheduler** | Cosine warm restart | Cosine warm restart | Cosine warm restart |
| **Test reconstruction loss (MAE)** | 0.07 | 0.04 | 0.03 |
| **Train codebook utilization** | 43.3% | 91.1% | 100% |
| **Test codebook utilization** | 38.9% | 88.4% | 100% |

**Selection criteria:** We chose QINCo as our final ECG tokenizer because it achieves the lowest reconstruction loss (0.03 vs 0.04 RVQ vs 0.07 VQ), a 12% improvement over standard RVQ and 53% over vanilla VQ. It achieves near-perfect codebook utilization (100.0%) vs 88.4% for RVQ and 38.9% for vanilla VQ, eliminating the codebook collapse problem.

### Supplementary Table 5 | ECG tokenizer codebook size sweep

| Codebook Size | Test reconstruction loss | Active codes |
|---|---|---|
| **128** | 0.04 | 100% |
| **256** | 0.03 | 100% |
| **512** | 0.03 | 100% |
| **1024** | 0.03 | 99.5% |

**Selection criteria:** We chose our current variant of the ECG tokenizer which has 8 quantizers and 512 codes since it has the best balance of test reconstruction loss and active code usage.

#### Supplementary Table 6 | Stage 2 training (Q-Former) hyperparameters.

| Parameter | Value | Search range |
|---|---|---|
| Bridge transformer layers | 10 | {6, 8, 10} |
| Hidden dimension | 768 | n/a |
| Attention heads | 8 | {8, 12} |
| Query tokens | 16 | {16, 32, 48} |
| Cross-attention frequency | Every 2 layers | {1, 2} |
| Codebooks kept | 8 | {1, 2, 4, 8} |
| Codebook size | 512 codes | n/a |
| Learning rate | 1.4e-4 | log-uniform [5e-5, 4e-4] |
| Weight decay | 0.01 | {0, 0.01, 0.02} |
| Optimizer | AdamW ($\beta_1$ = 0.9, $\beta_2$ = 0.999) | n/a |
| Batch size | 120 | {64, 96, 120} |
| Epochs | 10 | n/a |
| Precision | bfloat16 | n/a |

| | | |
|---|---|---|
| Gradient accumulation | 1 | n/a |
| Max text length | 128 tokens | n/a |
| Hard negative k | 3 | {1, 2, 3} |
| ETM warmup steps | 2,000 | n/a |
| ETG delay steps | 2,000 | n/a |
| ETG warmup to full weight | 4,000 steps | n/a |
| Focal γ (positive) | 2.0 | {1.0, 1.5, 2.0} |
| Focal γ (negative) | 0.0 | {0.0, 0.25, 0.5} |
| Text bank size | 2,048 | n/a |
| Bank refresh interval | 256 steps | n/a |
| Bank negatives per ECG | 5 | n/a |
| Max positives per ECG | 10 | n/a |

**Selection criterion:** Lowest loss

### Supplementary Table 7 | LoRA Hyperparameter Sweep

| **Hyperparameter** | **Values Tested** | **Selected** |
|---|---|---|

| | | |
|---|---|---|
| LoRA rank (r) | 16, 32, 48 | 32 |
| LoRA alpha | 32, 64, 96 | 64 |
| LoRA dropout | fixed | 0.05 |
| LoRA top-k layers | 8, 12, 16 | 12 |
| LoRA target modules | q, k, v, o_proj | q, k, v, o_proj |
| LLM learning rate | 2e-5 to 8e-5 (log-uniform) | 5e-5 |
| Bridge learning rate | 2e-4 to 1.2e-3 (log-uniform) | 5e-4 |
| LLM weight decay | 0, 1e-6, 1e-5 | 1e-5 |
| Bridge weight decay | 0, 1e-6, 1e-5 | 1e-5 |
| Scheduler | cosine_with_warmup, linear_warmup | cosine_with_warmup |
| Warmup fraction | 0, 0.03, 0.08 | 0.26 |
| Instruction dropout | 0.0, 0.1, 0.2 | 0.2 |
| Bridge dropout | 0.05, 0.1, 0.15 | 0.1 |
| Query tokens | 24, 32, 40 | 32 |
| Batch size | 8, 16 | 16 |
| Gradient accumulation steps | 16, 24, 32 | 16 |

Selection criterion: LLM-as-a-judge composite score on MHI internal validation set.

#### Supplementary Table 8 | ECG Interpretation Diagnoses

| Pathological | Limit | Normal |
|---|---|---|
| 2nd degree AV block - mobitz 2 | 1st degree AV block | ~~Monomorph~~ |
| 2nd degree AV block - mobitz 1 | ~~Early repolarization~~ | Sinus rhythm |
| Acute MI | Left anterior fascicular block | Regular rhythm |
| Acute pericarditis | Left axis deviation | |
| Afib | Left posterior fascicular block | |
| Atrial flutter | Nonspecific intraventricular conduction delay | |
| Atrial paced | ~~QRS complex negative in III~~ | |
| Atrial tachycardia (>= 100 BPM) | ~~R complex in V5-V6~~ | |
| LV pacing | ~~R/S ratio in V1-V2 >1~~ | |
| Bi-atrial enlargement | ~~RV1 + SV6 > 11 mm~~ | |
| Brugada | ~~RaVL > 11 mm~~ | |
| Delta wave | Right axis deviation | |
| Ectopic atrial rhythm (< 100 BPM) | Right bundle branch block | |
| Irregularly irregular | ~~ST upslopping~~ | |
| Junctional rhythm | ~~T wave inversion (inferior - II, III, aVF)~~ | |
| Left atrial enlargement | ~~T wave inversion (septal- V1-V2)~~ | |
| Left bundle branch block | U wave | |

| | |
|---|---|
| **Left ventricular hypertrophy** | ~~**qRS in V5-V6-I, aVL**~~ |
| **Prolonged QT** | ~~**rSR in V1-V2**~~ |
| **Q wave (anterior - V3-V4)** | |
| **Q wave (inferior - II, III, aVF)** | |
| **Q wave (lateral- I, aVL, V5-V6)** | |
| **Q wave (septal- V1-V2)** | |
| **Q wave (posterior - V7-V9)** | |
| ~~**QS complex in V1-V2-V3**~~ | |
| **Right atrial enlargement** | |
| **Right ventricular hypertrophy** | |
| **Right superior axis** | |
| **ST depression (inferior - II, III, aVF)** | |
| **ST depression (lateral - I, aVL, V5-V6)** | |
| ~~**ST depression (septal- V1-V2)**~~ | |
| ~~**ST depression (anterior - V3-V4)**~~ | |
| ~~**ST downslopping**~~ | |
| **ST elevation (anterior - V3-V4)** | |
| **ST elevation (inferior - II, III, aVF)** | |
| **ST elevation (lateral - I, aVL, V5-V6)** | |
| **ST elevation (posterior - V7-V8-V9)** | |
| **ST elevation (septal - V1-V2)** | |
| ~~**SV1 + RV5 or RV6 > 35 mm**~~ | |

| |
|---|
| **Supraventricular tachycardia** |
| ~~**T wave inversion (lateral -I, aVL, V5-V6)**~~ |
| ~~**T wave inversion (anterior - V3-V4)**~~ |
| **Third Degree AV Block** |
| **Ventricular paced** |
| **Ventricular Rhythm** |
| **Ventricular tachycardia** |
| **Wolff-Parkinson-White (Pre-excitation syndrome)** |

**Legend:** Strikethrough: Original labels removed as part of our evaluation to minimize overlap between diagnoses and collinearity.

#### Supplementary Table 9 | Linear vs EfficientNet probing AUROC across diagnostic categories

| **Category** | **Linear Probing** | **EfficientNet** |
|---|---|---|
| Overall | 0.79 (0.79-0.79) | 0.96 (0.96–0.96) |
| Rhythm | 0.75 (0.74-0.75) | 0.98 (0.98–0.98) |
| Conduction | 0.81 (0.81-0.82) | 0.97 (0.97–0.97) |
| Chamber Enlargement | 0.80 (0.79-0.80) | 0.96 (0.95–0.96) |
| Pericarditis | 0.85 (0.84-0.85) | 0.99 (0.99–0.99) |
| Infarct/Ischemia | 0.76 (0.76-0.77) | 0.96 (0.95–0.96) |
| Other | 0.82 (0.81-0.82) | 0.95 (0.95–0.96) |

#### Supplementary Table 10 | Report Generation Baselines

| | **Tokenizer + GPT-2 (epoch 1)** | **Tokenizer + GPT-2 (epoch 20)** | **Tokenizer + Llama 3.2-1B** | **ECG-Byte (Llama 3.2-1B) (95% CI)** |
|---|---|---|---|---|
| ECG interpretation (macro-AUROC) | | | | |
| Overall | 0.70 (0.69-0.70) | 0.78 (0.77-0.79) | 0.72 (0.71-0.73) | 0.50 (0.50-0.50) |
| Rhythm | 0.73 (0.72-0.75) | 0.81 (0.79-0.83) | 0.75 (0.73-0.77) | 0.50 (0.50-0.51) |
| Conduction | 0.70 (0.69-0.72) | 0.78 (0.76-0.80) | 0.70 (0.68-0.72) | 0.50 (0.50-0.50) |
| Chamber Enlargement | 0.61 (0.58-0.65) | 0.77 (0.73-0.80) | 0.70 (0.66-0.74) | 0.50 (0.50-0.50) |
| Pericarditis | 0.61 (0.55-0.68) | 0.81 (0.75-0.87)* | 0.69 (0.63-0.76) | 0.50 (0.50-0.52) |
| Infarct/Ischemia | 0.62 (0.61-0.63) | 0.76 (0.75-0.78) | 0.65 (0.64-0.67) | 0.50 (0.50-0.51) |
| Other | 0.73 (0.72-0.74) | 0.90 (0.89-0.90) | 0.76 (0.75-0.77) | 0.50 (0.50-0.50) |
| LLM metrics | | | | |
| ROUGE-1 | 0.51 | 0.66 | 0.56 | 0.37 |
| ROUGE-L | 0.50 | 0.66 | 0.55 | 0.31 |
| BLEU-1 | 0.52 | 0.66 | 0.57 | 0.23 |
| BLEU-4 | 0.27 | 0.45 | 0.32 | 0.05 |
| METEOR | 0.48 | 0.64 | 0.54 | 0.23 |

#### Supplementary Table 11 | Clinically curated hard-negative families for ECG-text matching

Families are grouped by selection mechanism: mutually exclusive groups (any non-matching member is a valid negative), targeted anchor-negative pairs (specific confusable conditions preferentially sampled when the anchor is the positive match), and yes/no counterparts (automatic answer-flip for binary QA).

| Mechanism | Family | Anchor or group | Negatives | Rationale |
|---|---|---|---|---|
| **Mutually exclusive** | Primary rhythm | Any rhythm label | All other members: sinus rhythm, atrial fibrillation, atrial flutter, atrial tachycardia, ectopic atrial rhythm, junctional rhythm, SVT, ventricular rhythm, ventricular tachycardia | One dominant rhythm mechanism per ECG |
| | Rhythm regularity | Any regularity label | All other members: regular, regularly irregular, irregularly irregular | Single categorical descriptor |
| | QRS axis | Any axis label | All other members: left axis deviation, right axis deviation, right superior axis | Frontal QRS axis occupies one quadrant |
| | Bundle branch block | LBBB or RBBB | The opposite laterality | Opposite QRS morphologies in the same leads |

| | | | | |
|---|---|---|---|---|
| | AV block degree | Any AV block grade | All other grades: 1st degree, 2nd degree Mobitz I, 2nd degree Mobitz II, 3rd degree | Block graded on a single severity scale |
| **Targeted anchor-negative (wide QRS)** | Wide QRS differential | LV pacing | Ventricular paced, ventricular rhythm, VT, LBBB | All produce wide QRS (>120 ms) with LBBB-like morphology; lead-specific criteria required to distinguish |
| | | Ventricular paced | LV pacing, ventricular rhythm, VT, LBBB | Pacemaker spikes may be absent on digital ECGs; paced QRS mimics LBBB |
| | | Ventricular rhythm | AFib, junctional rhythm, ventricular paced, LV pacing, VT | AIVR vs. slow VT vs. paced rhythm, all wide and regular |
| | | Ventricular tachycardia | Ventricular rhythm, AFib, SVT, ventricular paced | Wide-complex tachycardia differential (VT vs. SVT with aberrancy vs. pre-excited AFib) |
| **Targeted anchor-negative (ST polarity)** | Septal V1-V2 | ST elevation | ST depression (same leads) | Same territory, opposite deflection |
| | Anterior V3-V4 | ST elevation | ST depression (same leads) | Anterior changes separated by <1 mm in early presentations |

| | | | | |
|---|---|---|---|---|
| | Inferior II, III, aVF | ST elevation | ST depression (same leads) | Reciprocal changes in acute MI mimic primary ST depression |
| | Lateral I, aVL, V5-V6 | ST elevation | ST depression (same leads) | Subtle lateral ST deviations easily confused |
| | Posterior V7-V9 | ST elevation | ST depression (septal V1-V2), ST depression (anterior V3-V4) | Posterior STEMI presents as anterior/septal ST depression (mirror pattern) |
| **Targeted anchor-negative (AV block)** | AV block severity | Third-degree AV block | 1st degree, Mobitz I, Mobitz II, junctional rhythm | High-degree block mimics complete block when escape rate approximates atrial rate |
| **Targeted anchor-negative (repolarization)** | Repolarization mimics | U wave | Prolonged QT, low voltage, T-wave inversion (lateral), T-wave inversion (anterior) | Low-amplitude U waves confused with prolonged or bifid T waves and noise on low-voltage recordings |
| **Targeted anchor-negative (artifact)** | Artifact mimics | Lead misplacement | LV pacing, ventricular paced, ventricular rhythm | Limb lead reversal or precordial misplacement simulates wide-complex rhythms, paced morphology, or axis deviation |

| | | | | |
|---|---|---|---|---|
| **Yes/No counterpart** | Binary QA flip | Any binary diagnostic question (e.g., "Is there AFib? Yes") | The polarity-flipped counterpart (same question, opposite answer) | Prevents the model from exploiting text-only patterns; forces attention to the ECG signal |

#### Supplementary Table 12 | Q-Former pre-training convergence metrics

| Metric | Train | Validation |
|---|---|---|
| **Contrastive Recall@1** | 100% | 57.5% |
| **Contrastive Recall@5** | 100% | 87.8% |
| **ETG Next-Token Accuracy** | 90.2% | 87.6% |
| **ETM Accuracy** | 64.6% | 51.6% |
| **ETM AUROC** | 0.79 | 0.65 |
| **LT** | 0.01 | – |

#### Supplementary Table 13 | Numeric tolerance thresholds

| Parameter | Tolerance | Rationale |
|---|---|---|
| Heart rate | ±5 bpm | Standard ECG measurement variability |
| PR interval | ±20 ms | Inter-reader measurement variability |
| QT interval | ±20 ms | Waveform onset/offset definition variability |
| QTc interval | ±20 ms | Corrected interval measurement tolerance |
| LVEF | ±5% | Echocardiographic estimation variability |

### Supplementary Table 14 | External validation cohort characteristics

[a]Harvard-Emory reports are machine-generated by the Marquette 12SL analysis software (version 24), presented in lowercase, comma-separated format, which differs from the cardiologist-authored reports in the training data.

[b]Race/ethnicity data were available only for EchoNext; percentages are reported at the patient level.

[c]ECG type was classified by the validated BERT-based 77-condition classifier. This classifier applies only to interpretation cohorts; endpoint cohorts used linked echocardiographic data directly.

[d]Clinical endpoints and clinical settings apply only to endpoint cohorts (MIMIC-LVEF and EchoNext). Blank cells indicate that the variable was not applicable to the cohort.

| | **CLSA** | **Harvard-Emory** | **MIMIC-LVEF** | **EchoNext** |
|---|---|---|---|---|
| | **Interpretation Cohorts** | | **Clinical Endpoint Cohorts** | |
| **Cohort composition** | | | | |
| Source | CLSA baseline | Harvard-Emory | MIMIC-IV-ECG | EchoNext test set |
| Source population | Community-dwelling Canadian adults aged 45-85 years | Patients undergoing clinical ECG at MGH (Boston, MA) and EUH (Atlanta, GA), 1980-2022 | | Patients referred for electrocardiography at CUIMC (New York, NY) |
| Evaluation scope | Interpretation, classification | Interpretation, classification | LVEF prediction | SHD detection, LVEF prediction |
| Ground truth (interpretation) | Cardiologist-authored free-text reports | Machine-generated free-text reports[a] | | |
| Ground truth (classification/endpoints) | BERT-derived 77-class labels | BERT-derived 77-class labels | Linked echocardiography (LVEF) | Linked echocardiography (SHD+LVEF) |
| ECGs | 5,079 | 5,100 | 5,134 | 5,442 |

| | | | | |
|---|---|---|---|---|
| evaluated, n | | | | |
| Unique patients, n | 5,079 | 5,100 | 1,146 | 5,442 |
| **Demographics** | | | | |
| Age, year, mean (s.d.) | 65.3 (10.7) | 63.1 (16.9) | 59.3 (14.0) | 62.5 (16.4) |
| Male, n (%) | 2,867 (56.4%) | 2,750 (53.9%) | - | |
| Female, n (%) | 2,212 (43.6%) | 2,347 (46.0%) | - | |
| Race/ethnicity[b] | | | - | |
| **ECG type[c]** | | | | |
| Pathological, n (%) | 3,931 (77.4) | 4,339 (85.1) | - | |
| Borderline, n (%) | 834 (16.4) | 511 (10.0) | - | |
| Normal, n (%) | 314 (6.2) | 250 (4.9) | - | |
| **Clinical endpoints[d]** | | | | |
| LVEF available, n | - | - | 5,134 | 4,827 |
| LVEF, %, mean (s.d.) | - | - | 52.4 (14.2) | 53.5 (13.5) |
| Severely reduced (< 30%) | - | - | 485 (9.4) | 428 (8.9) |
| Moderately reduced (30-44%) | - | - | 807 (15.7) | 478 (9.9) |
| Mildly reduced (45-54%) | - | - | 887 (17.3) | 588 (12.2) |
| Normal (≥55%) | - | - | 2,955 (57.6) | 3,333 (69.0) |
| SHD available, n | - | - | - | 5,442 |

| | | | | |
|---|---|---|---|---|
| SHD present, n (%) | - | - | - | 2,318 (42.6) |
| SHD absent, n (%) | - | - | - | 3,124 (57.4) |
| Clinical setting[d] | | | | |
| Inpatient, n (%) | - | | | 2,203 (40.5) |
| Emergency, n (%) | - | | | 1,971 (36.2) |
| Outpatient, n (%) | - | | | 1,059 (19.5) |
| Procedural, n (%) | - | | | 209 (3.8) |

#### Supplementary Table 15 | QA prompt category distribution across external validation cohorts

Values are n (% of total QA pairs within each cohort). Blank cells indicate that the prompt category was not applicable to the cohort. Prompt categories were restricted to available ground truth: interpretation cohorts (CLSA, Harvard-Emory) were evaluated on report-based tasks; endpoint cohorts (MIMIC-LVEF, EchoNext) were evaluated on linked echocardiographic outcomes only. Up to four prompts were sampled per ECG per generation pass, selected by priority weight[a]. Categories with low prevalence (conduction disorders, chamber enlargement) received few QA pairs because higher-priority categories exhausted the per-ECG budget.

[a]Priority weights for prompt selection are defined in Supplementary Methods S6. QA, question-answer; LVEF, left ventricular ejection fraction.

| **Prompt category** | **CLSA** | **Harvard-Emory** | **MIMIC-LVEF** | **EchoNext** |
|---|---|---|---|---|
| **Interpretation** | 5,076 (25.0%) | 5,100 (25.0%) | – | – |
| **JSON interpretation** | 5,065 (25.0%) | 5,100 (25.0%) | – | – |
| **Classification** | 5,075 (25.0%) | 5,100 (25.0%) | – | – |
| **Rhythm disorders** | 3,794 (18.7%) | 3,738 (18.3%) | – | – |
| **Ischemia/infarction** | 1,254 (6.2%) | 1,210 (5.9%) | – | – |

| | | | | |
|---|---|---|---|---|
| **Conduction Disorders** | 10 (0.0%) | 138 (0.7%) | – | – |
| **Other findings** | 8 (0.0%) | 7 (0.0%) | – | – |
| **Chamber enlargements** | 7 (0.0%) | 7 (0.0%) | – | – |
| **LVEF** | – | – | 5,134 (100%) | 4,827 (47.0%) |
| **Structural heart disease** | – | – | – | 5,442 (53.0%) |
| **Total** | 20,289 | 20,400 | 5,134 | 10,269 |

#### Supplementary Table 16 | DeepECG-Tok vs DeepECG-SL & DeepECG-SSL AUROC

| **Category** | **DeepECG-Tok** | **DeepECG-SL** | **DeepECG-SSL** |
|---|---|---|---|
| Overall | 0.96 (0.96–0.96) | 0.94 (0.94–0.94) | 0.93 (0.93–0.94) |
| Rhythm | 0.98 (0.98–0.98) | 0.96 (0.96–0.96) | 0.96 (0.96–0.96) |
| Conduction | 0.97 (0.97–0.97) | 0.96 (0.96–0.97) | 0.96 (0.96–0.97) |
| Chamber Enlargement | 0.96 (0.95–0.96) | 0.92 (0.90–0.93) | 0.91 (0.90–0.92) |
| Pericarditis | 0.99 (0.99–0.99) | 0.97 (0.95–0.98) | 0.96 (0.94–0.98) |
| Infarct/Ischemia | 0.96 (0.95–0.96) | 0.92 (0.92–0.93) | 0.92 (0.91–0.92) |
| Other | 0.95 (0.95–0.96) | 0.91 (0.91–0.91) | 0.90 (0.89–0.90) |

#### Supplementary Table 17 | DeepECG-Tok EfficientNet vs ECG-GPT on PTBXL

| Label | n | Source | AUROC |
|---|---|---|---|
| AF | 1,514 | DeepECG-Tok | 0.99 (0.99-1.00) |

| | | | |
|---|---|---|---|
| | | ECG-GPT | 0.96 (0.95-0.96) |
| ST | 826 | DeepECG-Tok | 0.99 (0.99-0.99) |
| | | ECG-GPT | 0.95 (0.94-0.96) |
| SB | 637 | DeepECG-Tok | 0.93 (0.93-0.94) |
| | | ECG-GPT | 0.91 (0.90-0.92) |
| LBBB | 613 | DeepECG-Tok | 0.99 (0.98-0.99) |
| | | ECG-GPT | 0.97 (0.96-0.98) |
| RBBB | 1,658 | DeepECG-Tok | 0.94 (0.94-0.95) |
| | | ECG-GPT | 0.98 (0.98-0.99) |
| AVb | 823 | DeepECG-Tok | 0.96 (0.95-0.96) |
| | | ECG-GPT | 0.84 (0.82-0.85) |

#### Supplementary Table 18 | Cardiologist adjudication results (n = 500, per-category κ)

| **Task category** | **n** | **κ (95% CI)** |
|---|---|---|
| Free-text ECG interpretation | 90 | 0.83 (0.75–0.91) |
| Normal vs abnormal classification | 70 | 0.77 (0.68–0.86) |
| Rhythm abnormalities | 60 | 0.86 (0.76–0.95) |
| Infarction / ischaemia | 45 | 0.87 (0.76–0.99) |
| Conduction abnormalities | 35 | 0.69 (0.57–0.82) |
| Chamber enlargement | 25 | 0.72 (0.57–0.87) |
| Other diagnostic findings | 30 | 0.83 (0.69–0.97) |

| | | |
|---|---|---|
| Acute coronary syndrome severity | 30 | 0.80 (0.66–0.94) |
| Culprit-artery localisation | 20 | 0.82 (0.65–0.99) |
| Left ventricular ejection fraction | 40 | 0.77 (0.65–0.89) |
| Structural heart disease | 30 | 0.88 (0.74–1.00) |
| Incident atrial fibrillation risk | 25 | 0.88 (0.73–1.00) |
| **Overall (weighted)** | **500** | **0.81 (0.78–0.85)** |

**Abbreviations:** *κ, weighted Cohen's kappa between the automated LLM-as-a-Judge score and blinded cardiologist adjudication; CI, confidence interval.*

#### Supplementary Table 19 | Stage 3 internal per-category judge scores

| Prompt category | n | Score (95% CI) |
|---|---|---|
| Overall (category-weighted) | 49,776 | 0.71 (0.68-0.73) |
| Free-text interpretation | 10,000 | 0.63 (0.63-0.64) |
| JSON interpretation | 9,595 | 0.76 (0.76-0.77) |
| Classification | 8,343 | 0.52 (0.52-0.53) |
| Rhythm | 6,058 | 0.81 (0.80-0.81) |
| AFib risk | 3,121 | 0.63 (0.62-0.65) |
| Structural heart disease | 3,039 | 0.67 (0.65-0.69) |
| LVEF | 2,656 | 0.82 (0.81-0.83) |
| Ischemia/infarction | 2,597 | 0.71 (0.69-0.72) |
| Conduction | 1,553 | 0.73 (0.71-0.75) |
| Other diagnoses | 1,283 | 0.70 (0.68-0.72) |
| ACS severity | 1,021 | 0.59 (0.57-0.62) |
| Chamber enlargement | 275 | 0.86 (0.82-0.89) |

| | | |
|---|---|---|
| Culprit artery | 185 | 0.58 (0.55-0.61) |
| ECG interval | 16 | 0.56 (0.31-0.81) |
| Random findings | 14 | 0.80 (0.65-0.95) |
| Pericarditis | 10 | 1.00 (1.00-1.00) |
| T-wave localization | 6 | 0.57 (0.37-0.78) |
| QRS-axis localization | 3 | 1.00 (1.00-1.00) |
| Urgency assessment | 1 | 0.00 (0.00-0.00) |
| Q-wave localization | — | not evaluated (0 scored examples) |
| ST-segment localization | — | not evaluated (0 scored examples) |

Categories with n < 20 (ECG interval, random findings, pericarditis, T-wave localization, QRS-axis localization, urgency assessment) are shown for completeness; their confidence intervals are uninformative. Q-wave and ST-segment localization had no scored examples in the internal test set.

### Supplementary Table 20 | DeepECG-Tok outputs

| | **Prompt** | **Reference (ground truth)** | **Model output** | **Score** | **Judge rationale** |
|---|---|---|---|---|---|
| Chamber Enlargement [category score 0.86 (0.82-0.89)] | | | | | |
| Good | Can you assess for chamber abnormalities? | Left atrial enlargement; Left ventricular hypertrophy | Left atrial enlargement; Left ventricular hypertrophy | 1.00 | Exact match |
| Bad | Is there atrial enlargement? | Yes - Left atrial enlargement | No - no atrial enlargement | 0.00 | Binary answer mismatch (yes/no disagreement) |
| Rhythm disorders [category score 0.81 (0.80-0.81)] | | | | | |
| Good | Can you identify the cardiac rhythm? | Yes - Atrial tachycardia (>= 100 BPM); | Yes - Atrial tachycardia (>= 100 BPM); | 1.00 | Binary answer and descriptive details match |

| | | | | | |
|---|---|---|---|---|---|
| | | Ventricular Rhythm (HR: 129.0 bpm); Irregularly irregular rhythm; Atrial Fibrillation; Premature ventricular complex | Irregularly irregular rhythm (HR: 129.0 bpm); Atrial Fibrillation; Premature ventricular complex | | (1.00) |
| Bad | What is the rhythm in this ECG? | Atrial tachycardia (>= 100 BPM); Ventricular Rhythm (HR: 115.2 bpm); Irregularly irregular rhythm; Atrial Fibrillation; Premature ventricular complex | Sinus rhythm (HR: 65.0 bpm); Regular rhythm; Premature ventricular complex | 0.00 | No match |
| Conduction Disorders [category score 0.73 (0.71-0.75)] | | | | | |
| Good | Are there any fascicular blocks? | Yes - Left anterior fascicular block; Right bundle branch block; Left axis deviation | Yes - Left anterior fascicular block; Right bundle branch block; Left axis deviation | 1.00 | Exact match |
| Bad | Can you assess the conduction system? | 2nd degree AV block - mobitz 2; Left axis deviation; 1st degree AV block; Nonspecific intraventricular conduction delay | Ventricular paced | 0.00 | No match |
| Ischemia/infarction [category score 0.71 (0.69-0.72)] | | | | | |
| Good | Can you identify any signs of myocardial injury? | Yes - ST depression (lateral leads - I, avL, V5-V6); ST depression (inferior leads- II, | Yes - ST depression (lateral leads - I, avL, V5-V6); ST depression (inferior leads- II, | 1.00 | Binary answer and descriptive details match (1.00) |

| | | | | | |
|---|---|---|---|---|---|
| | | III, aVF); ST depression (anterior leads - V3-V4); ST depression in inferior - II, III, aVF; ST depression in anterior - V3-V4; ST depression in lateral - I, avL, V5-V6; T wave inversion in inferior - II, III, aVF; T wave inversion in lateral -I, aVL, V5-V6; T wave inversion in anterior - V3-V4; T wave inversion in septal- V1-V2 | III, aVF); ST depression (anterior leads - V3-V4); ST depression in inferior - II, III, AVF; ST depression in anterior - V3-v4; ST depression in septal- V1-V2; T wave inversion in lateral -I, aVL, V5-v6 | | |
| Bad | What do the ST segments show? | Previous septal (V1-V2) Myocardial Infarction; ST elevation (anterior - V3-V4); Previous inferior wall MI; Previous anterior (V3-V4) Myocardial Infarction; ST elevation (lateral leads- I, aVL, V5-V6); ST depression (lateral leads - I, avL, V5-V6); ST elevation (inferior leads - II, III, aVF) | Previous anterior (V3-V4) Myocardial Infarction | 0.00 | No match |
| Other findings [category score 0.70 (0.68-0.72)] | | | | | |

| | | | | | |
|---|---|---|---|---|---|
| Good | Are the T waves normal in morphology? | No - T wave inversions present in inferior - II, III, aVF; lateral -I, aVL, V5-V6; anterior - V3-V4; septal- V1-V2 | No - T wave inversions present in inferior - II, III, aVF; lateral -I, aVL, V5-V6; anterior - V3-V4; septal- V1-V2 | 1.00 | Exact match |
| Bad | What other findings are present? | T wave inversion (septal- V1-V2); T wave inversion (inferior - II, III, aVF); T wave inversion (anterior - V3-V4); T wave inversion (lateral - I, aVL, V5-V6) | Diffuse ST segment downslopping; Monomorph QRS complexes | 0.00 | No match |
| Classification [category score 0.52 (0.52-0.53)] | | | | | |
| Good | Is urgent action needed for this ECG? | Yes - there are pathological findings; Pathological findings: Previous anterior (V3-V4) Myocardial Infarction; Previous lateral (V5, V6, I, aVL) Myocardial Infarction; T wave inversion (anterior - V3-V4); T wave inversion (lateral - I, aVL, V5-V6) | Yes - there are pathological findings; Pathological findings: Previous anterior (V3-V4) Myocardial Infarction; T wave inversion (lateral - I, aVL, V5-V6) | 1.00 | Binary answer and descriptive details match (1.00) |
| Bad | Does this ECG require follow-up? | Yes - ECG shows significant abnormalities; Pathological findings: Wolff-Parkinson-White | Normal ECG; No significant abnormalities detected | 0.00 | Binary answer mismatch (yes/no disagreement) |

| | | | | | |
|---|---|---|---|---|---|
| | | (Pre-excitation syndrome); Delta wave; Left ventricular hypertrophy; ST depression (lateral leads - I, avL, V5-V6); ST depression (inferior leads- II, III, aVF) | | | |
| JSON interpretation [category score 0.76 (0.76-0.77)] | | | | | |
| Good | Output JSON ONLY with keys: RHYTHM, CONDUCTION, CHAMBER_ENLARGEMENT, INFARCT_ISCHEMIA, PERICARDITIS, heart_rate_bpm, ecg_classification. Values must be lists of present findings (omit missing categories). | {"RHYTHM":["Bradycardia","Sinusal","Regular","Premature ventricular complex"],"CONDUCTION":["Left anterior fascicular block","1st degree AV block"],"CHAMBER_ENLARGEMENT":["Left ventricular hypertrophy"],"INFARCT_ISCHEMIA":["Q wave_septal- V1-V2","Q wave_anterior_V3-V4","ST depression_lateral_I_avL_V5-V6"],"heart_rate_bpm":56,"ecg_classification":"pathological"} | {"RHYTHM":["Bradycardia","Sinusal","Regular"],"CONDUCTION":["Left anterior fascicular block","1st degree AV block"],"CHAMBER_ENLARGEMENT":["Left ventricular hypertrophy"],"INFARCT_ISCHEMIA":["Q wave_septal- V1-V2","ST depression_lateral_I_avL_V5-V6"],"heart_rate_bpm":57,"ecg_classification":"pathological"} | 0.94 | Schema valid: False, Fields: 6, Extra keys: 0 |
| Bad | Provide structured JSON output for this ECG | {"RHYTHM":["Bradycardia","Sinusal","Regular"],"CONDUCTION":[" | {"RHYTHM":["Bradycardia","Sinusal","Regular"],"CONDUCTION":[ | 0.00 | Invalid JSON format |

| | | | | | |
|---|---|---|---|---|---|
| | analysis using keys RHYTHM, CONDUCTION, CHAMBER_ENLARGEMENT, INFARCT_ISCHEMIA, PERICARDITIS, heart_rate_bpm, ecg_classification. | Left anterior fascicular block","Right bundle branch block","Left axis deviation"],"CHAMBER_ENLARGEMENT":["Left ventricular hypertrophy"],"INFARCT_ISCHEMIA":["ST elevation_anterior_V3-V4","ST elevation_lateral_I_aVL_V5-V6","ST depression_lateral_I_avL_V5-V6","Acute MI","ST elevation_septal_V1-V2","ST elevation_inferior_II_III_aVF","ST depression_inferior_II_III_aVF"],"heart_rate_bpm":54,"ecg_classification":"pathological"} | "Left anterior fascicular block","Right bundle branch block","Left axis deviation","Prolonged QT"],"CHAMBER_ENLARGEMENT":["Left ventricular hypertrophy"],"INFARCT_ISCHEMIA":["ST depression_lateral_I_avL_V5-V6","ST depression_inferior_II_III_aVF"],"heart_rate_bpm":54," | | |
| Free-text interpretation [category score 0.63 (0.63-0.64)] | | | | | |
| Good | What does this electrocardiogram reveal? | *** ACUTE MI *** - *** AGE AND SEX SPECIFIC ECG ANALYSIS *** Normal sinus rhythm ST elevation (HR: 73.0 bpm); possibility of inferior injury or acute infarct ** ** ACUTE MI / STEMI ** ** | *** ACUTE MI *** - Normal sinus rhythm ST elevation (HR: 73.0 bpm); possibility of inferior injury or acute infarction ** ** ACUTE MI / STEMI ** ** Consider right ventricular involvement, | 1.00 | F1: 1.00, Penalty: 0.0 |

| | | | | | |
|---|---|---|---|---|---|
| | | Consider right ventricular involvement, acute inferior infarct Abnormal ECG | acute inferior infarct Abnormal ECG | | |
| Bad | Please describe the ECG findings. | *** ACUTE STEMI (anterior - V3-V4, septal - V1-V2, inferior - II, III, aVF, lateral - I, aVL, V5-V6) *** - Pacer detection suspended due to external noise-REVIEW ADVISED; HR: 92.0 bpm;*** CONSIDER ACUTE ST ELEVATION MI ;Sinus arrhythmia with PVC(s);Short PR interval;Left axis deviation;RBBB with left anterior fascicular block; LATERAL INFARCT - POSSIBLY ACUTE ***;Inferior and ant/septal ST elevation. CONSIDER ACUTE INFARCT;Abnormal ECG | Ventricular pacing (HR: 91.0 bpm);Pacemaker rhythm - no further analysis;Abnormal ECG | 0.00 | F1: 0.000, Penalty: 0.0 |

#### Supplementary Table 21 | DeepECG-Tok vs ECG-GPT on PTBXL (LLM AUROCs)

| **Label** | **Tokenizer + GPT-2 (epoch 20)** | **DeepECG-Tok (MedGemma-4B)** | **ECG-GPT** |
|---|---|---|---|

| | | | |
|---|---|---|---|
| AF | 0.92 (0.92-0.93) | 0.92 (0.91-0.93) | 0.96 (0.95-0.96) |
| ST | 0.95 (0.94-0.96) | 0.96 (0.94-0.97) | 0.95 (0.94-0.96) |
| SB | 0.90 (0.89-0.91) | 0.91 (0.89-0.92) | 0.91 (0.90-0.92) |
| LBBB | 0.96 (0.95-0.97) | 0.95 (0.93-0.96) | 0.97 (0.96-0.98) |
| RBBB | 0.96 (0.95-0.97) | 0.96 (0.95-0.98) | 0.98 (0.98-0.99) |
| AVb | 0.66 (0.64-0.68) | 0.66 (0.64-0.69) | 0.84 (0.82-0.85) |

### Supplementary Table 22 | Binary classification performance for LVEF across internal and external cohorts

| | **Task** | **MHI (internal) n = 2,656** | **MIMIC-LVEF (external) n = 5,134** | **EchoNext (external) n = 4,827** |
|---|---|---|---|---|
| Prevalence | LVEF≤40 | 20.3% | 20.5% | 15.8% |
| | LVEF<50 | 31.2% | 32.0% | 22.7% |
| Sensitivity | LVEF≤40 | 0.66 (0.62-0.70) | 0.46 (0.43-0.49) | 0.48 (0.45-0.52) |
| | LVEF<50 | 0.57 (0.54-0.60) | 0.39 (0.36-0.41) | 0.42 (0.39-0.45) |
| Specificity | LVEF≤40 | 0.82 (0.81-0.84) | 0.90 (0.89-0.91) | 0.89 (0.88-0.90) |
| | LVEF<50 | 0.85 (0.84-0.87) | 0.92 (0.91-0.93) | 0.91 (0.90-0.91) |
| PPV | LVEF≤40 | 0.48 (0.45-0.52) | 0.54 (0.51-0.57) | 0.46 (0.43-0.49) |
| | LVEF<50 | 0.64 (0.60-0.67) | 0.69 (0.66-0.72) | 0.57 (0.53-0.60) |
| NPV | LVEF≤40 | 0.90 (0.89-0.92) | 0.87 (0.86-0.88) | 0.90 (0.89-0.91) |
| | LVEF<50 | 0.82 (0.80-0.83) | 0.76 (0.75-0.77) | 0.84 (0.83-0.85) |
| AUROC | LVEF≤40 | 0.80 (0.77-0.82) | 0.76 (0.75-0.78) | 0.74 (0.72-0.75) |

| | | | | |
|---|---|---|---|---|
| | LVEF<50 | 0.77 (0.75-0.79) | 0.73 (0.72-0.74) | 0.71 (0.69-0.73) |
| MAE | | 8.41% (8.11-8.73) | 10.21% (9.98-10.44) | 10.22% (9.97-10.47) |
| Pearson r | | 0.56 (0.52-0.59) | 0.46 (0.43-0.48) | 0.41 (0.38-0.44) |
| ICC | | 0.68 (0.64-0.71) | 0.60 (0.58-0.63) | 0.51 (0.47-0.54) |

#### Supplementary Table 23 | Signal dependency analysis: Per-category judge score

| Category | n | Normal | Zeroed | Shuffled |
|---|---|---|---|---|
| interpretation | 243 | 0.31 | 0.02 | 0.14 |
| classification | 243 | 0.40 | 0.08 | 0.24 |
| category_rhythm | 133 | 0.49 | 0.26 | 0.28 |
| category_infarct_ischemia | 110 | 0.58 | 0.41 | 0.43 |
| category_conduction | 10 | 0.76 | 0.53 | 0.56 |
| json_interpretation | 243 | 0.37 | 0.36 | 0.28 |

#### Supplementary Table 24 | Signal dependency analysis: representative model outputs under altered ECG input conditions

| Task (ground truth) | Condition | Model output |
|---|---|---|
| Interpretation (Atrial fibrillation) | Normal | Atrial fibrillation Abnormal ECG (HR: 80) |
| | Zeroed | Analysis impossible; no QRS detectable |
| | Shuffled | Ventricular paced rhythm, intrinsic P waves |

| | | |
|---|---|---|
| Classification (Normal ECG) | Normal | No, ECG is normal |
| | Zeroed | No, ECG is normal |
| | Shuffled | Yes, irregularly irregular; atrial fibrillation |
| Ischemia/infarction (Previous inferior wall MI) | Normal | Previous inferior wall MI |
| | Zeroed | No ST changes |
| | Shuffled | Previous septal (V1-V2) MI |
| Other findings (No ectopic beats) | Normal | No, no ectopic beats present |
| | Zeroed | No, no ectopic beats present |
| | Shuffled | No, no ectopic beats present |

Representative examples from the signal dependency analysis (n = 996 QA pairs). Each row shows the model output and LLM-as-a-judge score for a single question-answer pair evaluated under three ECG input conditions: the correct preprocessed ECG (normal), an all-zeros tensor (zeroed), and a randomly selected ECG from a different patient (shuffled). Examples were selected to illustrate the range of observed behaviors, from complete signal dependence (interpretation, ischemia/infarction) through partial dependence (classification, where zeroed input produces the correct default but shuffled input overrides it) to text-prior dominance (other findings, where all conditions produce identical outputs). MI, myocardial infarction; HR, heart rate; QRS, ventricular depolarization complex.

#### Supplementary Table 25 | External per-category classification AUROC (frozen-tokenizer EfficientNetV2 multi-label classification)

| Category | Internal (MHI) | CLSA | Harvard-Emory |
|---|---|---|---|
| Overall | 0.96 (0.96-0.96) | 0.88 (0.87-0.91) | 0.90 (0.89-0.91) |
| Rhythm | 0.98 (0.98-0.98) | 0.95 (0.94-0.96) | 0.97 (0.96-0.97) |
| Conduction | 0.97 (0.97-0.97) | 0.94 (0.91-0.96) | 0.94 (0.93-0.95) |
| Infarct/Ischemia | 0.96 (0.95-0.96) | 0.85 (0.82-0.87) | 0.88 (0.85-0.90) |
| Chamber Enlargement | 0.96 (0.95-0.96) | 0.84 (0.77-0.89) | 0.84 (0.82-0.86) |

| | | | |
|---|---|---|---|
| Other | 0.95 (0.95-0.96) | 0.82 (0.80-0.84) | 0.82 (0.80-0.84) |

Macro-averaged AUROC for frozen-tokenizer (EfficientNetV2) multi-label classification by diagnostic category, for the internal MHI test set (n = 416,467) and the two external cohorts (CLSA, n = 5,079; Harvard-Emory, n = 5,100). Values are AUROC (95% CI) from 1,000 bootstrap resamples. External cohorts were compared with internal by two-sided bootstrap difference test; overall AUROC differed from internal for both cohorts (P < 0.001), with Harvard-Emory rhythm the exception (P = 0.22). Abbreviations: AUROC - area under the receiver operating characteristic curve; CLSA - Canadian Longitudinal Study on Aging; MHI - Montreal Heart Institute.

### Supplementary Table 26 | External per-category judge scores (CLSA, Harvard-Emory)

#### CLSA

| **Prompt category** | **LLM-as-a-judge score (95% CI)** | **P** |
|---|---|---|
| Overall (n = 20,289) | 0.53 (0.53–0.54) | <0.001 |
| Free-text interpretation (n = 5,076) | 0.64 (0.63–0.64) | 0.72 (ns) |
| JSON interpretation (n = 5,065) | 0.42 (0.41–0.42) | <0.001 |
| Classification (n = 5,075) | 0.45 (0.44–0.46) | <0.001 |
| Rhythm (n = 3,794) | 0.60 (0.58–0.61) | <0.001 |
| Ischemia / infarction (n = 1,254) | 0.69 (0.67–0.71) | 0.46 (ns) |
| Conduction (n = 10) | 0.53 (0.32–0.73) | 0.07 (ns) |
| Chamber enlargement (n = 7) | 0.93 (0.79–1.00) | 0.65 (ns) |
| Other (n = 8) | 0.48 (0.19–0.79) | 0.13 (ns) |

#### Harvard-Emory

| **Prompt category** | **LLM-as-a-judge score (95% CI)** | **P** |
|---|---|---|
| Overall (n = 20,400) | 0.50 (0.49–0.50) | <0.001 |
| Free-text interpretation (n = 5,100) | 0.54 (0.54–0.55) | <0.001 |

| | | |
|---|---|---|
| JSON interpretation (n = 5,100) | 0.38 (0.37–0.39) | <0.001 |
| Classification (n = 5,100) | 0.46 (0.45–0.47) | <0.001 |
| Rhythm (n = 3,738) | 0.57 (0.56–0.58) | <0.001 |
| Ischemia / infarction (n = 1,210) | 0.66 (0.64–0.68) | 0.01 |
| Conduction (n = 138) | 0.72 (0.66–0.78) | 0.93 (ns) |
| Chamber enlargement (n = 7) | 0.86 (0.57–1.00) | 0.79 (ns) |
| Other (n = 7) | 0.46 (0.14–0.86) | 0.15 (ns) |

#### Supplementary Table 27 | Stage 1 fairness analysis: equalized-odds disparities by sex and age.

Per-attribute ΔTPR and ΔFPR (maximum minus minimum across subgroup levels) with 95% bootstrap confidence intervals (1,000 resamples, seed = 42), reported for the pooled internal test set and per cohort (MHI, MIMIC-IV). TPR and FPR were micro-averaged across all 77 diagnostic labels within each subgroup. Sex levels: male, female. Age bands: below 55, 55 to 75, above 75 years. Values below 0.10 denote acceptable disparity. ΔTPR, true-positive-rate disparity; ΔFPR, false-positive-rate disparity; TPR, true-positive rate; FPR, false-positive rate; MHI, Montreal Heart Institute; MIMIC, Medical Information Mart for Intensive Care.

| **Cohort** | **Attribute** | **ΔTPR (95% CI)** | **ΔFPR (95% CI)** | **Below 0.10** |
|---|---|---|---|---|
| Pooled internal | Sex (Male vs Female) | 0.009 (0.008–0.011) | 0.018 (0.018–0.019) | Yes |
| Pooled internal | Age (<55 / 55–75 / >75) | 0.024 (0.022–0.025) | 0.047 (0.046–0.048) | Yes |
| MHI | Sex (Male vs Female) | 0.001 (0.000–0.003) | 0.021 (0.020–0.022) | Yes |
| MHI | Age (<55 / 55–75 / >75) | 0.025 (0.023–0.027) | 0.030 (0.029–0.031) | Yes |
| MIMIC-IV | Sex (Male vs Female) | 0.007 (0.005–0.009) | 0.016 (0.015–0.017) | Yes |
| MIMIC-IV | Age (<55 / 55–75 / >75) | 0.008 (0.006–0.011) | 0.071 (0.070–0.072) | Yes |

#### Supplementary Table 28 | Stage 3 fairness analysis: LLM-as-a-judge weighted-composite score by subgroup, with subgroup-difference tests.

Weighted-composite judge scores with 95% bootstrap confidence intervals (1,000 resamples, seed = 42) for each sex and age subgroup, reported for the pooled internal test set and per cohort (MHI, MIMIC-IV). Subgroup-difference tests report the composite difference (95% CI), rank-biserial effect size, and Holm-Bonferroni-adjusted two-sided Mann-Whitney U P value against the reference group (female sex; age below 55 years). Equalized odds was undefined for free-text generation; composite-score parity was used at Stage 3 in its place. Significance: *P < 0.05, **P < 0.01, ***P < 0.001; ns, not significant. CI, confidence interval; MHI, Montreal Heart Institute; MIMIC, Medical Information Mart for Intensive Care.

(a) Weighted-composite LLM-as-a-judge score by subgroup

| Cohort | Subgroup | n (QA pairs) | Composite (95% CI) |
|---|---|---|---|
| Pooled internal | Overall | 49,776 | 0.71 (0.68–0.73) |
| | Male | 29,257 | 0.70 (0.67–0.73) |
| | Female | 20,129 | 0.71 (0.68–0.74) |
| | Age <55 | 9,467 | 0.72 (0.68–0.76) |
| | Age 55–75 | 24,584 | 0.72 (0.69–0.76) |
| | Age >75 | 15,226 | 0.68 (0.64–0.72) |
| MHI | Overall | 24,995 | 0.75 (0.73–0.76) |
| | Male | 16,168 | 0.75 (0.74–0.76) |
| | Female | 8,437 | 0.72 (0.70–0.74) |
| | Age <55 | 4,254 | 0.71 (0.68–0.74) |
| | Age 55–75 | 13,178 | 0.73 (0.70–0.76) |
| | Age >75 | 7,173 | 0.74 (0.71–0.77) |
| MIMIC-IV | Overall | 24,781 | 0.70 (0.68–0.73) |
| | Male | 13,089 | 0.69 (0.66–0.73) |
| | Female | 11,692 | 0.69 (0.65–0.74) |
| | Age <55 | 5,213 | 0.73 (0.69–0.77) |

| | | | |
|---|---|---|---|
| | Age 55–75 | 11,406 | 0.72 (0.68–0.77) |
| | Age >75 | 8,053 | 0.65 (0.60–0.70) |

(b) Subgroup-difference tests (reference: Female; Age <55)

| Cohort | Comparison | ΔComposite (95% CI) | Rank-biserial | Mann–Whitney U P (Holm-adjusted) | Sig |
|---|---|---|---|---|---|
| Pooled internal | Male vs Female | −0.01 (−0.06, +0.04) | +0.03 | <0.001 | *** |
| | Age 55–75 vs <55 | +0.00 (−0.05, +0.05) | +0.04 | <0.001 | *** |
| | Age >75 vs <55 | −0.04 (−0.10, +0.01) | +0.09 | <0.001 | *** |
| MHI | Male vs Female | +0.03 (+0.00, +0.05) | +0.02 | 0.017 | * |
| | Age 55–75 vs <55 | +0.02 (−0.02, +0.06) | +0.00 | 0.655 | ns |
| | Age >75 vs <55 | +0.03 (−0.01, +0.07) | +0.03 | 0.036 | * |
| MIMIC-IV | Male vs Female | +0.00 (−0.05, +0.06) | +0.04 | <0.001 | *** |
| | Age 55–75 vs <55 | −0.01 (−0.07, +0.05) | +0.08 | <0.001 | *** |
| | Age >75 vs <55 | −0.08 (−0.14, −0.02) | +0.15 | <0.001 | *** |

#### Supplementary Table 29 | Natural language generation metrics on the held-out internal interpretation test set

| Model | ROUGE-1 | ROUGE-2 | ROUGE-L | BLEU-1 | BLEU-4 | METEOR | n |
|---|---|---|---|---|---|---|---|
| MedGemma 4B-IT 1.0 + QFormer | 0.71 | 0.57 | 0.69 | 0.65 | 0.51 | 0.71 | 10,000 |
| MedGemma 4B-IT 1.5 + QFormer | 0.69 | 0.54 | 0.67 | 0.64 | 0.48 | 0.70 | 10,000 |

#### Supplementary Table 30 | Category Weights and Judge Assignments

| Category | Weight | Judge Type | Output Format |
|---|---|---|---|
| Interpretation | 1.5 | LLM adjudication | Free text |
| JSON interpretation | 1.2 | Deterministic | Structured JSON |
| ACS severity | 1.2 | Deterministic | 6-class categorical |
| Ischemia/infarction | 1.2 | LLM adjudication | Free text |
| Classification | 1.0 | Deterministic | Categorical |
| LVEF | 1.0 | Deterministic | Percentage |
| Rhythm | 1.0 | LLM adjudication | Free text |
| Conduction | 1.0 | LLM adjudication | Free text |
| Chamber enlargement | 1.0 | LLM adjudication | Free text |
| AFib risk | 1.0 | Deterministic | Binary |

| | | | |
|---|---|---|---|
| Urgency assessment | 1.0 | Deterministic | Ordinal categorical |
| Culprit artery | 1.0 | Deterministic | Multi-class categorical |
| Structural heart disease | 1.0 | Deterministic | Binary |
| ST localization | 1.0 | LLM adjudication | Anatomical regions |
| T-wave localization | 1.0 | LLM adjudication | Anatomical regions |
| Other findings | 1.0 | LLM adjudication | Free text |
| ECG interval | 0.8 | Deterministic | Numeric values |

### Supplementary Figures

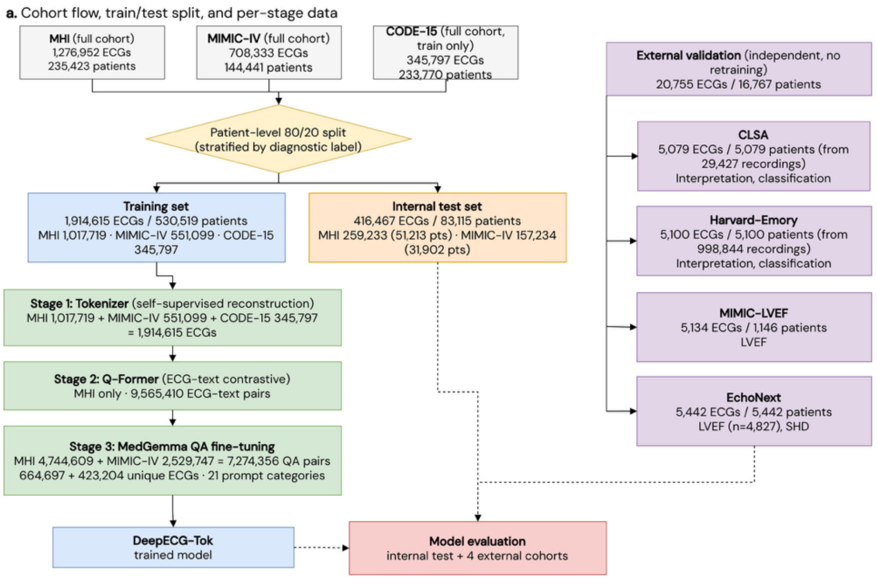

**b.** ECG content type by dataset

| Dataset | Pathological | Borderline | Normal | Denominator |
|---|---|---|---|---|
| MHI | 557,300 (83.8%) | 58,020 (8.7%) | 49,377 (7.4%) | QA set: 664,697 ECGs |
| MIMIC-IV | 343,098 (81.1%) | 54,002 (12.8%) | 26,104 (6.2%) | QA set: 423,204 ECGs |
| CODE-15 | 61.1% | n/a | 38.9% | full cohort: 345,797 (binary flag) |

**c.** Stage 3 QA prompt-category distribution (n = 7,274,356)

| Prompt category | QA pairs | % |
|---|---|---|
| Interpretation | 1,491,307 | 20.5 |
| JSON interpretation | 1,422,240 | 19.6 |
| Classification | 1,394,209 | 19.2 |
| Rhythm | 1,042,305 | 14.3 |
| AFib risk | 499,839 | 6.9 |
| Infarct / ischemia | 425,023 | 5.8 |
| Conduction | 285,167 | 3.9 |
| Structural heart disease | 254,460 | 3.5 |
| LVEF | 197,422 | 2.7 |
| Other | 184,398 | 2.5 |
| Chamber enlargement | 42,296 | 0.6 |
| ACS severity | 28,373 | 0.4 |
| Pericarditis | 1,590 | <0.1 |
| ECG interval | 1,516 | <0.1 |
| Random finding | 1,234 | <0.1 |
| QRS-axis localization | 1,152 | <0.1 |
| Culprit artery | 919 | <0.1 |
| T-wave localization | 489 | <0.1 |
| Q-wave localization | 329 | <0.1 |
| Urgency assessment | 70 | <0.1 |
| ST-depression localization | 18 | <0.1 |
| **Total** | **7,274,356** | **100** |

**Supplementary Fig. 1 | Study cohorts, data partitioning, and per-stage data usage across the DeepECG-Tok pipeline.** a, Cohort flow showing the three source datasets (MHI, 1,276,952 ECGs from 235,423 patients; MIMIC-IV, 708,333 ECGs from 144,441 patients; and CODE-15, 345,797 ECGs from 233,770 patients, training only), partitioned by a patient-level 80/20 split stratified by diagnostic label into a training set (1,914,615 ECGs from 530,519 patients) and an internal test set (416,467 ECGs from 83,115 patients). Per-stage data usage is shown for Stage 1 tokenizer pre-training (self-supervised reconstruction; 1,914,615 ECGs), Stage 2 Q-Former ECG-text contrastive alignment (MHI only; 9,565,410 ECG-text pairs) and Stage 3 MedGemma question-answer (QA) fine-tuning (7,274,356 QA pairs derived from 664,697 MHI and 423,204 MIMIC-IV unique ECGs across 21 prompt categories). The trained model was evaluated on the internal test set and four independent external cohorts with no retraining (20,755 ECGs from 16,767 patients): CLSA, Harvard-Emory, MIMIC-LVEF and EchoNext. b, Distribution of ECG content type (pathological, borderline and normal) by dataset. c, Distribution of the 7,274,356 Stage 3 QA pairs across the 21 prompt categories. MHI, Montreal Heart Institute; MIMIC, Medical Information Mart for Intensive Care; CLSA, Canadian Longitudinal Study on Aging; LVEF, left ventricular ejection fraction; SHD, structural heart disease; QA, question-answer.

Vanilla VQ — Sample 1 (Overall MAE: 0.0850)
Dataset: MHI | Report: Normal sinus rhythm, normal ECG

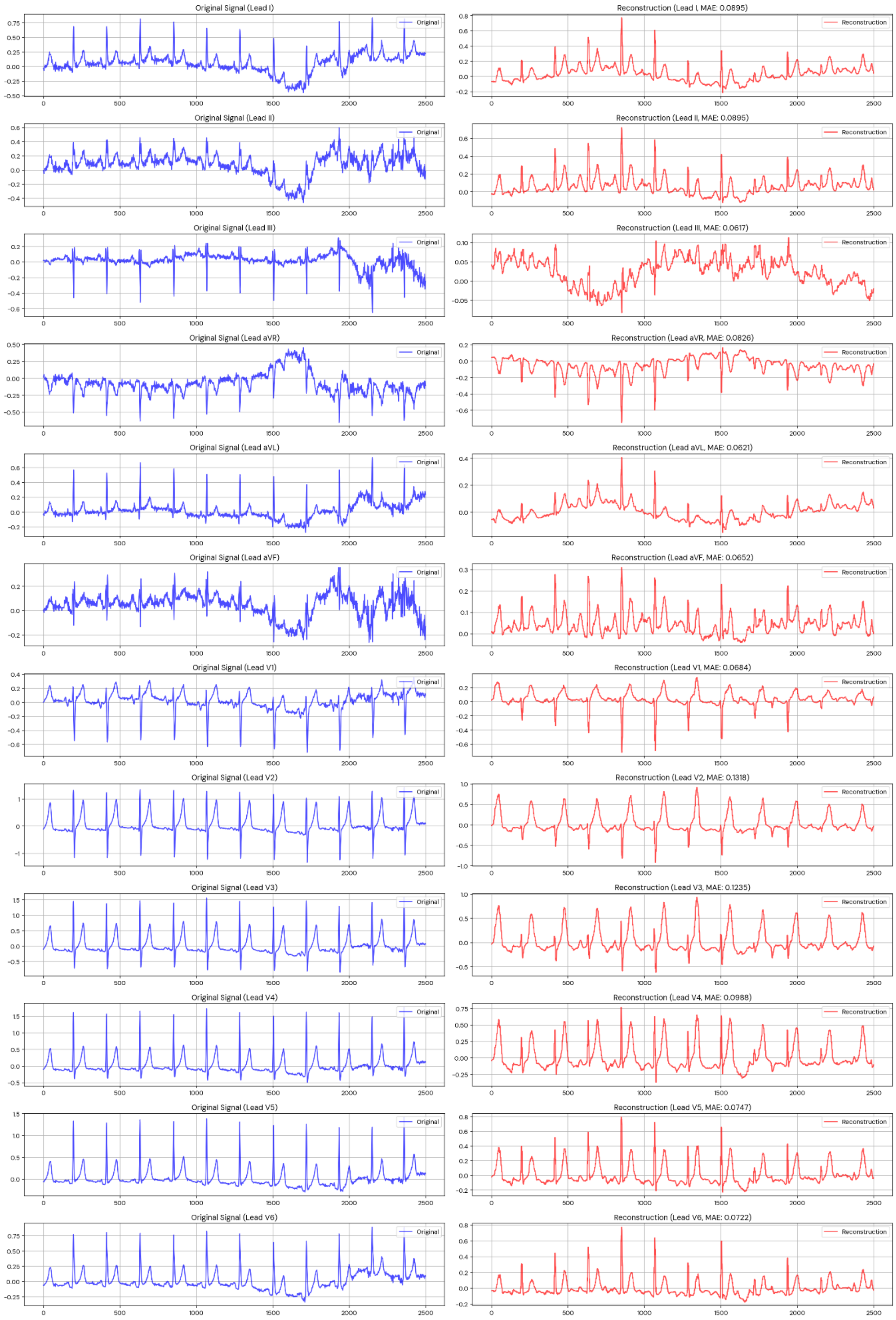

**Supplementary Fig. 2 | Reconstruction fidelity of the Vanilla VQ tokenizer (1 codebook, 512 codes).** Twelve-lead ECG reconstruction for a representative test sample from the MHI cohort (clinical report: "Normal sinus rhythm, normal ECG"). The left column shows the preprocessed waveform for each lead; the right column shows the reconstructed signal after vector quantization with a single codebook. Per-lead MAE is shown in each reconstruction panel title. Test-set overall MAE: 0.07 (sample shown: 0.09).

RVQ — Sample 1 (Overall MAE: 0.0434)
Dataset: MHI | Report: Normal sinus rhythm, normal ECG

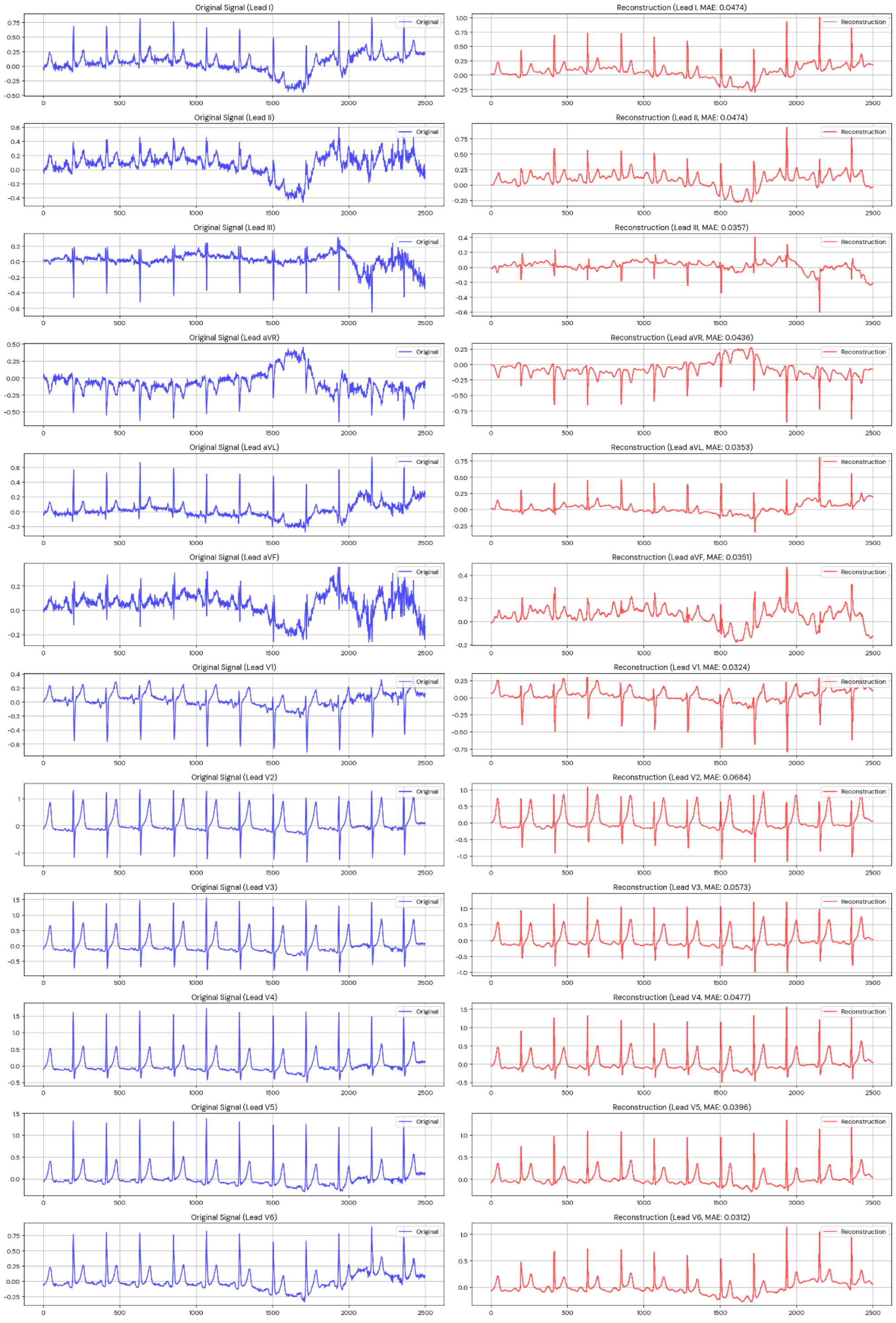

**Supplementary Fig. 3 | Reconstruction fidelity of the RVQ tokenizer (8 codebooks, 512 codes each).** Twelve-lead ECG reconstruction for a representative test sample from the MHI cohort (clinical report: "Normal sinus rhythm, normal ECG"). The left column shows the preprocessed waveform for each lead; the right column shows the reconstructed signal after residual vector quantization with 8 sequential codebooks. Per-lead MAE is shown in each reconstruction panel title. Test-set overall MAE: 0.04 (sample shown: 0.04).

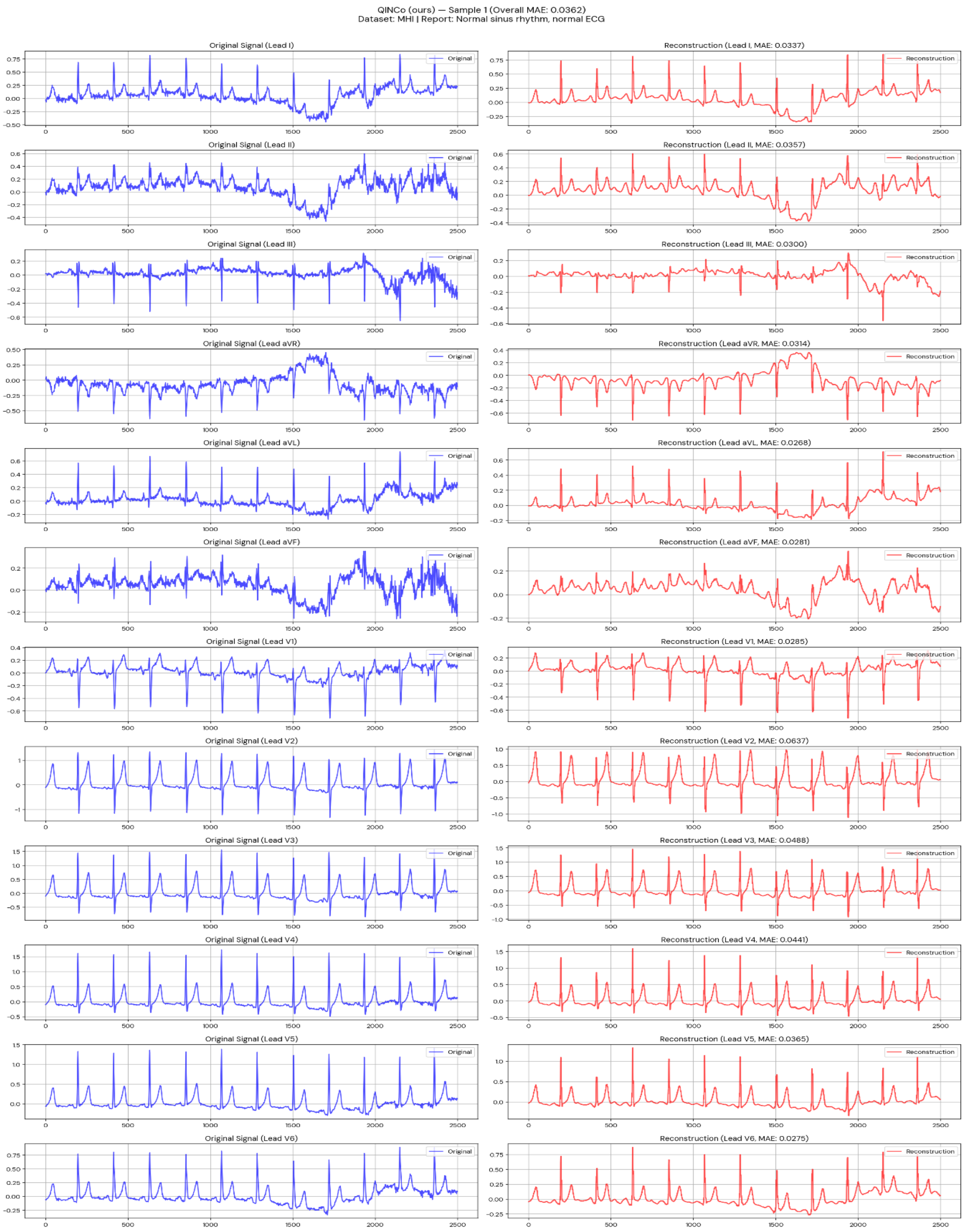
QINCo (ours) — Sample 1 (Overall MAE: 0.0362)
Dataset: MHI | Report: Normal sinus rhythm, normal ECG
Original Signal (Lead I)
Reconstruction (Lead I, MAE: 0.0337)
Original Signal (Lead II)
Reconstruction (Lead II, MAE: 0.0357)
Original Signal (Lead III)
Reconstruction (Lead III, MAE: 0.0300)
Original Signal (Lead aVR)
Reconstruction (Lead aVR, MAE: 0.0314)
Original Signal (Lead aVL)
Reconstruction (Lead aVL, MAE: 0.0268)
Original Signal (Lead aVF)
Reconstruction (Lead aVF, MAE: 0.0281)
Original Signal (Lead V1)
Reconstruction (Lead V1, MAE: 0.0285)
Original Signal (Lead V2)
Reconstruction (Lead V2, MAE: 0.0637)
Original Signal (Lead V3)
Reconstruction (Lead V3, MAE: 0.0488)
Original Signal (Lead V4)
Reconstruction (Lead V4, MAE: 0.0441)
Original Signal (Lead V5)
Reconstruction (Lead V5, MAE: 0.0365)
Original Signal (Lead V6)
Reconstruction (Lead V6, MAE: 0.0275)
Original
Reconstruction

**Supplementary Fig. 4 | Reconstruction fidelity of the QINCo tokenizer (8 adaptive codebooks, 512 codes each).** Twelve-lead ECG reconstruction for a representative test sample from the MHI cohort (clinical report: "Normal sinus rhythm, normal ECG"). The left column shows the preprocessed waveform for each lead; the right column shows the reconstructed signal after adaptive residual vector quantization (QINCo), where each codebook is conditioned on the evolving residual via an implicit neural network. Per-lead MAE is shown in each reconstruction panel title. Test-set overall MAE: 0.03 (sample shown: 0.04).
